# Evaluation of Clonal Hematopoiesis and Mosaic Loss of Y Chromosome in Cardiovascular Risk: an analysis in prospective studies

**DOI:** 10.1101/2024.01.15.24301313

**Authors:** S Fawaz, S Marti, M Dufossée, Y Pucheu, A Gaufroy, J Broitman, A Bidet, A Soumaré, G Munsch, C Tzourio, S Debette, DA Trégouët, C James, O Mansier, T Couffinhal

## Abstract

**Background:** Clonal hematopoiesis of indeterminate potential (CHIP) was initially linked to a twofold increase in atherothrombotic events. However, recent investigations have revealed a more nuanced picture, suggesting that CHIP may confer only a modest rise in Myocardial Infarction (MI) risk. This observed lower risk might be influenced by yet unidentified factors that modulate the pathological effects of CHIP. Mosaic loss of Y chromosome (mLOY), a common marker of clonal hematopoiesis in men, has emerged as a potential candidate for modulating cardiovascular risk associated with CHIP. In this study, we aimed to ascertain the risk linked to each somatic mutation or mLOY and explore whether mLOY could exert an influence on the cardiovascular risk associated with CHIP.

**Methods:** We conducted an examination for the presence of CHIP and mLOY using targeted high-throughput sequencing and digital PCR in a cohort of 446 individuals. Among them, 149 patients from the CHAth study had experienced a first myocardial infarction (MI) at the time of inclusion (MI(+) subjects), while 297 individuals from the Three-City cohort had no history of cardiovascular events (CVE) at the time of inclusion (MI(-) subjects). All subjects underwent thorough cardiovascular phenotyping, including a direct assessment of atherosclerotic burden. Our investigation aimed to determine whether mLOY could modulate inflammation, atherosclerosis burden, and atherothrombotic risk associated with CHIP.

**Results:** CHIP and mLOY were detected with a substantial prevalence (45.1% and 37.7%, respectively), and their occurrence was similar between MI(+) and MI(-) subjects. Notably, nearly 40% of CHIP(+) male subjects also exhibited mLOY. Interestingly, neither CHIP nor mLOY independently resulted in significant increases in plasma hsCRP levels, atherosclerotic burden, or MI incidence. Moreover, mLOY did not amplify or diminish inflammation, atherosclerosis, or MI incidence among CHIP(+) male subjects. Conversely, in MI(-) male subjects, CHIP heightened the risk of MI over a five-year period, particularly in those lacking mLOY.

**Conclusion:** Our study highlights the high prevalence of CHIP and mLOY in elderly individuals. Importantly, our results demonstrate that neither CHIP nor mLOY in isolation substantially contribute to inflammation, atherosclerosis, or MI incidence. Furthermore, we find that mLOY does not exert a significant influence on the modulation of inflammation, atherosclerosis burden, or atherothrombotic risk associated with CHIP. However, CHIP may accelerate the occurrence of MI, especially when unaccompanied by mLOY. These findings underscore the complexity of the interplay between CHIP, mLOY, and cardiovascular risk, suggesting that large-scale studies with thousands more patients may be necessary to elucidate subtle correlations.

## Introduction

Atherothrombosis is the main cause of death worldwide. Traditional cardiovascular risk factors (CVRF), such as diabetes, smoking, dyslipidemia, hypertension, explain 70 to 75% of cardiovascular events suffered by patients. However, a significant part of these events remains unexplained given that 25 to 30% of people without any evident cause can present an atherosclerotic cardiovascular event (CVE), whereas not all high-risk subjects (according to traditional CVFR) experience such an event (Berry et al., 2012).

Recently, clonal hematopoiesis of indeterminate potential (CHIP) has emerged as a potential new risk factor of cardiovascular diseases (Jaiswal et al., 2014). This condition results from the acquisition by a hematopoietic stem cell of somatic mutations in leukemia-driver genes, leading to clonal expansion of a population of hematopoietic cells without any clinical or biological sign of hematological malignancy. The definition of CHIP proposed to date, requires the detection of the mutation at a variant allele frequency (VAF) of more than 2%, representing a proportion of mutated cells of more than 4% (Steensma et al., 2015). The most commonly mutated genes in CHIP are *DNA Methyltransferase 3A* (*DNMT3A*) and *Ten Eleven Translocation 2* (*TET2*). In 2014, Jaiswal *et al* showed that CHIP was associated with a decreased survival mainly because of an increased atherothrombotic mortality. In particular, they observed a 2.0 fold-increased risk of myocardial infarction (MI) and a 2.6-fold increased risk of ischemic stroke (Jaiswal et al., 2014). In 2017, these data were confirmed, showing a 1.9-fold increased risk of coronary heart disease in the presence of CHIP, independently of traditional CVRF (Jaiswal et al., 2017). At the same time, a causative role of CHIP in inducing atherosclerosis has been demonstrated in animal models through the induction of a proinflammatory state (Fuster et al., 2017; Jaiswal et al., 2017). More recently, Kessler *et al* showed in 454 803 subjects from the UK Biobank that the association of CHIP with atherothrombotic events was restricted to high VAF clones (*ie* ≥10%) with a much lower risk than initially demonstrated (HR=1.11, Kessler et al., 2022). But even with these criteria, no association between CHIP and atherothrombotic events was found in a validation cohort of 173 585 subjects (Kessler et al., 2022), which has also been suggested in another work studying several hundred thousand subjects (Kar et al., 2022). Thus, the impact of CHIP on atherothrombosis in humans is not totally evident, possibly because of the existence of yet unidentified modulating factors that could potentiate or counteract the effect of CHIP. For example, the p.Asp358Ala variant of the IL6 receptor gene has been shown to decrease the atherothrombotic risk associated with CHIP (Bick et al., 2020; Vlasschaert et al., 2023a). However, the impact of other genetic variants on the cardiovascular risk associated with CHIP remains unknown.

Gain or loss of chromosomes in hematopoietic cells appears to be as frequent as the acquisition of somatic mutations during aging (Saiki et al., 2021). In particular, mosaic loss of chromosome Y (mLOY) has been shown to be frequent in male subjects without evidence of hematological malignancy (Wright et al., 2017; Zhou et al., 2016). mLOY was associated with cardiovascular diseases (Loftfield et al., 2018; Sano et al., 2022), and can be detected in a rather high proportion of subjects with CHIP (Ljungström et al., 2022; Zink et al., 2017). Finally, while *TET2* mutations promote inflammation and atherosclerosis in mouse models (Jaiswal et al., 2017), mLOY was shown to switch macrophages from a pro-inflammatory to a pro-fibrotic phenotype (Sano et al., 2022). Thus, because of their opposite effect on macrophages phenotype, mLOY could balance the effect of CHIP regarding the induction of inflammation and thus decrease the development of atherosclerosis and the resulting atherothrombotic risk. We thus hypothesized that mLOY could modulate the effect of CHIP in inducing inflammation, atherosclerosis and triggering atherothrombotic events.

In this study, we used sensitive technics to determine the prevalence of both CHIP and mLOY in 2 cohorts of subjects. We sought to determine whether CHIP and mLOY significantly increase the cardiovascular risk separately. We also searched to determine in humans whether mLOY could impact the effect of CHIP on inflammation, atherosclerosis burden or atherothrombotic risk.

## Methods

### Patients

For this study, we recruited 446 patients: 149 with a first MI and 297 without a MI or other CVE at inclusion. The 149 subjects with a MI were enrolled in the CHAth study between March 2019 and October 2021 (MI(+) subjects). The main eligibility criterion was suffering from a first MI of atherosclerosis origin after 75 years of age without evidence of hematological malignancy (Supplementary Figure S1). They were included 4+/-2 months after the acute event, in order to assess their basal inflammatory state. In the presence of any clinical sign or factor associated with inflammation, the appointment was reported in order not to skew biological and genetic data. Additionally, we ensured that the subjects had not been vaccinated against SARS-Cov2 within 15 days of enrollment. The study was approved by the institutional review board (IRDCB 2019-A02902-05), and registered (https://www.clinicaltrials.gov: NCT04581057). All participants gave written informed consent before inclusion in the study. Eligibility and exclusion criteria are more detailed in the Supplementary Methods.

As a control cohort, we selected subjects without any history of CVE at inclusion in the Three-City (3C) study cohort (MI(-) subjects, Supplementary Figure S1) (3C Study Group, 2003). The 3C study is a prospective study that enrolled 9294 subjects of 65 years or more who were selected upon electoral lists. These subjects were followed for several years (up to 12 years) to detect the development of dementia from a vascular origin. As such, they benefitted from a stringent cardiovascular follow-up, with adjudication of all cardiovascular events (in particular occurrence of MI). Among these subjects, we selected the 297 subjects who did not present any CVE before inclusion. Seventy-nine of them presented a MI during follow-up. The remaining 218 subjects had no atherothrombotic event during follow up, and were matched on age, sex and CVRF with those who had a MI during follow up (Supplementary Figure S1).

### Clinical and biological parameters measured at inclusion

In MI(+) subjects, a routine cardiovascular evaluation was performed at inclusion (between 2 and 7 month after MI occurrence). Subjects were asked about presence of dyspnea or angina, evaluated with NYHA and CCS scales. Information was obtained from medical records about a potential recurrence of a cardiovascular event since the index event (new MI, coronary revascularization, stroke, hospitalization for acute heart failure). Traditional cardiovascular risk factors were noted. Routine biological analyses comprising blood count, high-sensitive CRP (hsCRP), lipid profile, HbA1c were performed in the laboratory of the university hospital of Bordeaux on fresh samples.

For MI(-) subjects, data available at inclusion included hsCRP level, lipid profile and traditional CVRF. No blood count was available but none of the subjects developed cancer (including hematological malignancy) during follow-up suggesting that detectable somatic mutations were indicative of CHIP and not hematological malignancy.

### Measurement of atherosclerosis burden

For MI(+) subjects, a transthoracic echocardiography was performed at inclusion by trained cardiologists and ejection fraction calculated. Supra-aortic trunks ultrasonography with 3D-measurement of carotid atheroma volume was performed by trained physicians of the hospital University of Bordeaux, using a Philips iU22 probe equipped with a linear-3D volume convertor VL13-5 (Philips). Images were analyzed using the Vascular Plaque Quantification software on the QLAB 10.2 system (Philips). Carotid stenosis quantification was done with NASCET criteria. Functional ischemic testing was performed at the clinicians’ discretion. For MI(-) subjects, atherosclerosis burden was assessed at the inclusion by ultrasound echography recording detection of atherosclerotic plaques, atherosclerotic plaque numbers and intima-media thickness.

### Follow up of subjects

For MI(+) subjects, the follow-up was conducted with a standardized questionnaire previously validated in clinical trials (Lafitte et al., 2013). Recurrence of MI as well as any significant cardiovascular event (cardiovascular death, acute coronary syndrome, stroke or transient ischemic attack, congestive heart failure, secondary coronary revascularization, or peripheral vascular surgery) occurring between the initial event and the year after inclusion in the study were recorded. All medical records of participants who died, or who reported on the questionnaire that they had experienced cardiovascular symptoms between baseline and follow-up evaluations, were reviewed by one of the investigators, and the patient practitioners were contacted. For MI(-) subjects, a follow up was performed for up to 12 years. All cardiovascular events were recorded (including MI) with adjudication upon medical records by an expert committee (Supplementary Figure 1).

### Search for somatic mutations and mosaic loss of chromosome Y

For both MI(+) and MI(-) subjects, DNA was extracted from total leukocytes obtained at inclusion. The search for somatic mutations was carried out by the Laboratory of Hematology of the University Hospital of Bordeaux using the Next Generation Sequencing (NGS) panel designed for the diagnosis and follow-up of myeloid hematological malignancies. The genes tested are detailed in Supplementary Table S1.

Briefly, target sequences were captured using the SureSelect technology (Agilent). Sequencing was performed on a NextSeq 550Dx Instrument (Illumina technology) and analyzed using an in-house bioinformatics pipeline (see Supplementary Methods for more details). Variants interpretation was performed independently by two biologists according to criteria previously described (Luque Paz et al., 2021) and reported in Supplementary Table S2. According to the definition of CHIP, only mutations with a VAF ≥ 2% were retained. Although we did not used error-corrected sequencing, the high depth (2111X in median) coupled with bioinformatic tools and manual curing allowed us to reliably detect variants with a VAF≥1%.

The search of mLOY was performed thanks to an in-house droplet digital PCR technic using the following primers and probes:

- Primer-amel-Fwd : 5’- CCCCTGGGCACTGTAAAGAAT
- Primer-amel-Rev: 5’- CCAAGCATCAGAGCTTAAACTG
- Probe-amelX : 5’- HEX-CCAAATAAAGTGGTTTCTCAAGT-BHQ
- Probe-amelY: 5’- FAM-CTTGAGAAACATCTGGGATAAAG-BHQ.

Briefly, 75 ng of DNA was mixed with ddPCR supermix for Probes (no dUTP, Biorad), primers (0.9µM each) and probes (0.25µM each). The emulsion was prepared with the QX-100 (Biorad). The amplification program was as follows: 10-minutes denaturation at 95°C, followed by 40 cycles of 30 seconds at 94°C, 1 minute at 55°C, and inactivation of 10 minutes at 98°C. The number of droplets positive for amelX and amelY was determined on the QX-200 droplet reader (Biorad) using the Quantasoft software version 1.5 (Biorad). At least 10,000 droplets were analyzed in each well. We determined the background noise of our technique by analyzing the DNA of control subjects (men under 40 years old with a normal karyotype, as assessed by conventional cytogenetic studies). We observed that only a signal corresponding to 9% of cells with mLOY could be considered different from background noise. By analyzing a dilution series of control DNA, we demonstrated the reliability of our ddPCR assay in estimating the proportion of cells with mLOY and its ability to detect as low as 10% of cells with mLOY. Considering the background noise, we established our threshold for confirming the presence of mLOY at 9% of cells with mLOY.

### Statistical analyses

Univariate association analyses were conducted using Fisher exact or Chi-square test statistics for categorical variables. Analysis of variance was used for quantitative variables. Multivariate association analyses were performed using logistic or linear regression models as appropriate. Analyses were adjusted for age and sex (CHIP) or for age only (mLOY). We used raw values for all quantitative variables as they presented a normal distribution, except for CRP levels for which we analyzed log(CRP). Log-rank test and Cox statistical models were employed to assess the association of clinical/biological variables with the incidence of future cardiovascular events.

All analyses were conducted using either the RStudio software (Posit team (2023). RStudio: Integrated Development Environment for R. Posit Software, PBC, Boston, MA. URL http://www.posit.co/.) or the PRISM software (GraphPad Prism version 9.5.1).

## Results

### Patient’s characteristics

In this study, we aimed to decipher whether mLOY could alter the effect of CHIP in inducing a chronic inflammation that would favor the development of atherosclerosis and the incidence of atherothrombotic events. To answer this question, we analyzed 446 subjects from 2 prospective studies, 149 who presented a MI at inclusion and 297 who did not present any CVE before inclusion. The general characteristics of the 446 combined subjects are detailed in Table 1. Briefly, the median age was 76.4 years and 257 (57.6%) were males. Forty percent of them presented more than 2 CVRF. MI(+) subjects were older (due to the inclusion criteria), more frequently men, and presented a lower cardiovascular risk than MI(-) subjects (due to the initiation of treatment between the initial event and the inclusion).

**Table 1:** Characteristics of the population at the time of inclusion.

|  | All subjects<br>n = 446 | MI(+) subjects<br>n=149 | MI(-) subjects<br>n=297 | p-value |
| --- | --- | --- | --- | --- |
| Male, n (%) | 257 (57.6) | 98 (66) | 159 (54) | 0.015 |
| Median age, years (Q1;Q3) | 76.4 (71.9;80.9) | 82.0 (78.0;86.0) | 73.6 (70.6;77.8) | P < 10<sup>-4</sup> |
| **Cardiovascular risk factors** |  |  |  |  |
| BMI, kg/m<sup>2</sup> (Q1;Q3) | 25.5 (23.6;28.3) | 25.5 (23.6;28.5) | 25.5 (23.6;28.1) | 0.14 |
| Diabetes, n (%) | 94 (21.2) | 47 (32%) | 47 (16%) | p < 10<sup>-4</sup> |
| Hypertension, n (%) | 362 (82.8) | 107 (76.4) | 255 (85.9) | 0.020 |
| Total cholesterol, g/L (Q1;Q3) | 2.08 (1.72;2.38) | 1.45 (1.25;1.72) | 2.23 (2.00;2.47) | p < 10<sup>-4</sup> |
| LDL-c, g/L (Q1;Q3) | 1.25 (0.94;1.52) | 0.77 (0.61;1.03) | 1.39 (1.19;1.60) | p < 10<sup>-4</sup> |
| HDL-c, g/L (Q1;Q3) | 0.56 (0.46;0.66) | 0.47 (0.39;0.57) | 0.59 (0.50;0.68) | p < 10<sup>-4</sup> |
| Smoking, n (%) | 32 (7.2) | 6 (4.4) | 26 (8.7) | 0.117 |
| **Prevalence of CHIP and mLOY** |  |  |  |  |
| CHIP prevalence, n (%) | 201 (45.1) | 79 (53%) | 122 (41%) | 0.923 |
| Prevalence of CHIP with VAF ≥5% | 88 (19.7) | 30 (20.1) | 58 (19.5) | 0.069 |
| Subjects tested for mLOY, n | 220 | 97 | 123 | - |
| mLOY prevalence, n (%) | 83 (37.7) | 44 (45.4) | 39 (31.7) | 0.783 |
| CHIP(+) / mLOY(+) prevalence, n (%) | 39 (17.7) | 22 (22.7) | 17 (13.8) | 0.797 |

Data are expressed as numbers and frequency or median, first and third quartiles. For quantitative values, comparisons were made by linear regression of log values adjusted for age and sex. For qualitative parameters, comparisons were made by the fisher test. For each variable, results are expressed among patients with available value.

### CHIP and mLOY are detected as frequently in MI(+) and MI(-) subjects

Among the 446 subjects, at least one mutation with a VAF≥2% was detected in 201 persons (45.1%), defining CHIP(+) subjects (Table 1, Figure 1A). As previously described, *DNMT3A* and *TET2* were the 2 most frequently mutated genes. The other mutated genes were those previously described in CHIP (Figure 1B, Jaiswal et al., 2017, 2014; Zink et al., 2017) and the median VAF was 2% (Figure 1C). The mutational profile of CHIP(+) subjects is available in the Supplementary Tables S3 and S4 while the characteristics of CHIP(+) compared with CHIP(-) subjects are detailed in the Supplementary Tables S5 and S6. In this study, we considered subjects without any detectable mutation or with only mutations with a VAF below 2% as non-CHIP carriers (CHIP(-) subjects).

**Figure 1:**
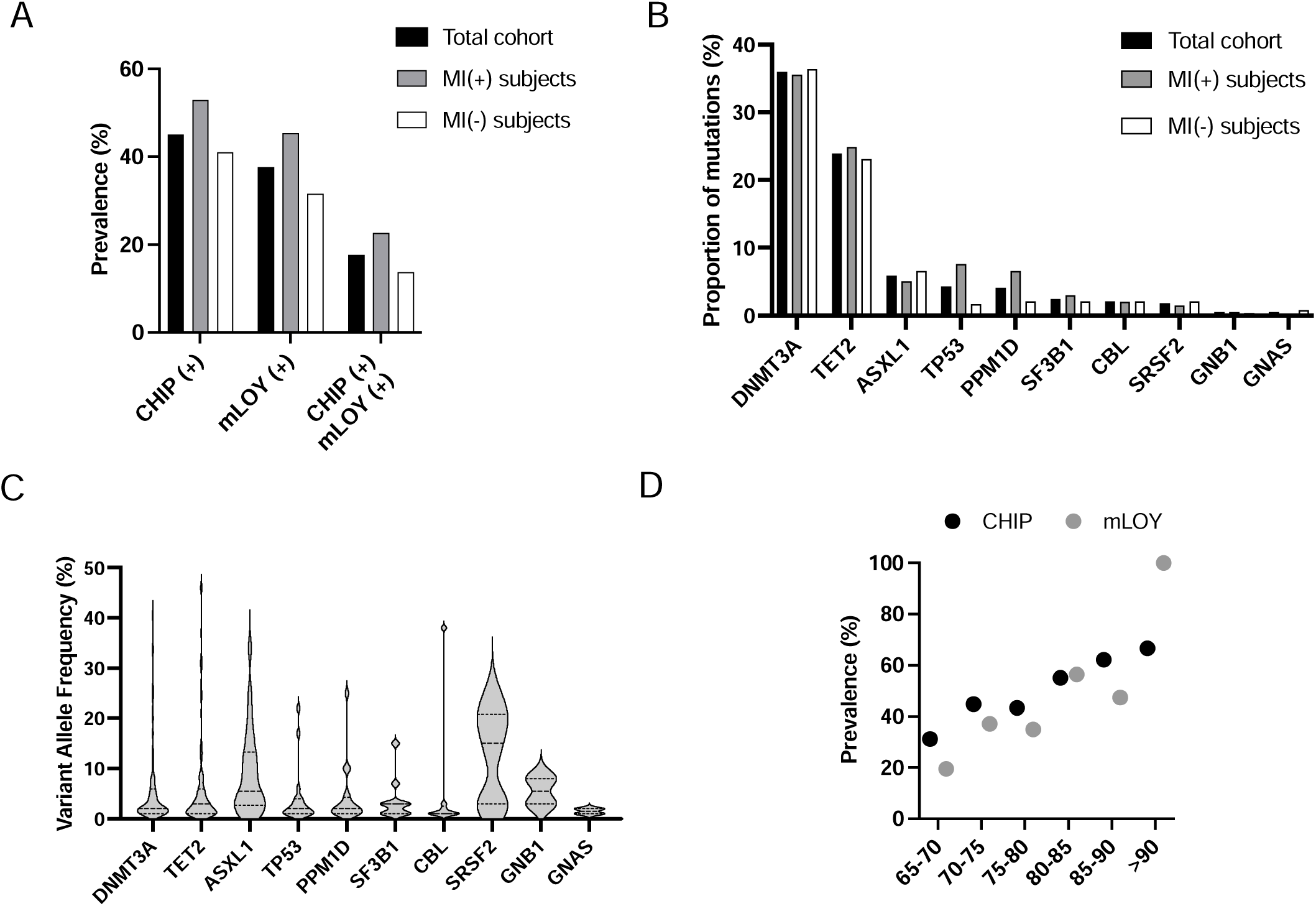
CHIP, mLOY and their combination are as frequent in MI(+) and MI(-) subjects. A: Prevalence of CHIP, mLOY and their combination in the total cohort of 449 subjects, in MI(+) as well as in MI(-) subjects. B: Mutational spectrum of CHIP expressed as the proportion of mutations detected in the indicated genes. C: VAF measured for the different mutations detected in the 449 subjects detected in the indicated genes. D: Prevalence of CHIP and mLOY depending on age in the total cohort (for CHIP) and in male subjects (for mLOY).

A mLOY was present in 83 (37.7%) male subjects (mLOY(+) subjects, Table 1, Figure 1A). The clinico-biological characteristics of mLOY(+) subjects compared with mLOY(-) subjects are detailed in the Supplementary Tables S5 and S6. The median proportion of cells with mLOY was 18% [12%;32%]. There was no significant association between CHIP and mLOY since 39 CHIP(+) subjects (39.8%) also carried a mLOY compared with 44 CHIP(-) subjects (36.1%, p=0.579). Finally, as previously demonstrated, CHIP(+) subjects were significantly older than CHIP(-) subjects (Supplementary Tables S5 and S6). We also observed a significant association between age and CHIP prevalence (p<0.001). Similarly, the prevalence of mLOY increased with age (p<0.001, Figure 1D).

We observed a similar frequency of CHIP and mLOY in MI(+) and MI(-) subjects (Figure 1A, Table 1). Besides, the association of CHIP with mLOY was as frequent in MI(+) and MI(-) subjects (22.7% and 13.8%, p=0.797). Of note, MI(+) and MI(-) subjects presented similar proportions of mutated genes (Figure 1B) and VAF (Supplementary Figure 2). Similar results were also observed when considering CHIP associated with important clones (*i.e.* VAF≥5%, Table 1), when analyzing only *DNMT3A* and *TET2* mutations (data not shown), or when making further adjustment on CVRF.

Altogether, these results suggest that CHIP and mLOY are very frequent but not associated with the existence of a history of MI, even when mLOY is associated with CHIP.

### Neither CHIP, mLOY, nor their combination highly increase basal inflammation or atherosclerotic burden

It was demonstrated that *TET2* mutations were associated with the induction of a pro-inflammatory phenotype of macrophages (Fuster et al., 2017). On the contrary, mLOY was shown to decrease the inflammatory phenotype of macrophages (Sano et al., 2022). Therefore, we searched to determine whether mLOY could counter the inflammatory state that would be associated with CHIP. In the total cohort, CHIP(+) and CHIP(-) subjects presented similar levels of hsCRP, as did mLOY(+) and mLOY(-) subjects (Table 2). Similarly, hsCRP levels were not different in CHIP(-)/mLOY(-), CHIP(+)/mLOY(-), CHIP(-)/mLOY(+) and CHIP(+)/mLOY(+) subjects. The impact of CHIP and mLOY on hsCRP levels, either alone or in combination, was comparable in MI(+) or MI(-) subjects (Table 2). This was also true when restricting the analysis to *DNMT3A* and *TET2* mutated CHIP(+) subjects (Supplementary Table S7), or when adjusting further on CVRF. Finally, subjects with important CHIP or mLOY clones did not present higher levels of hsCRP (Supplementary Table S8).

**Table 2:**
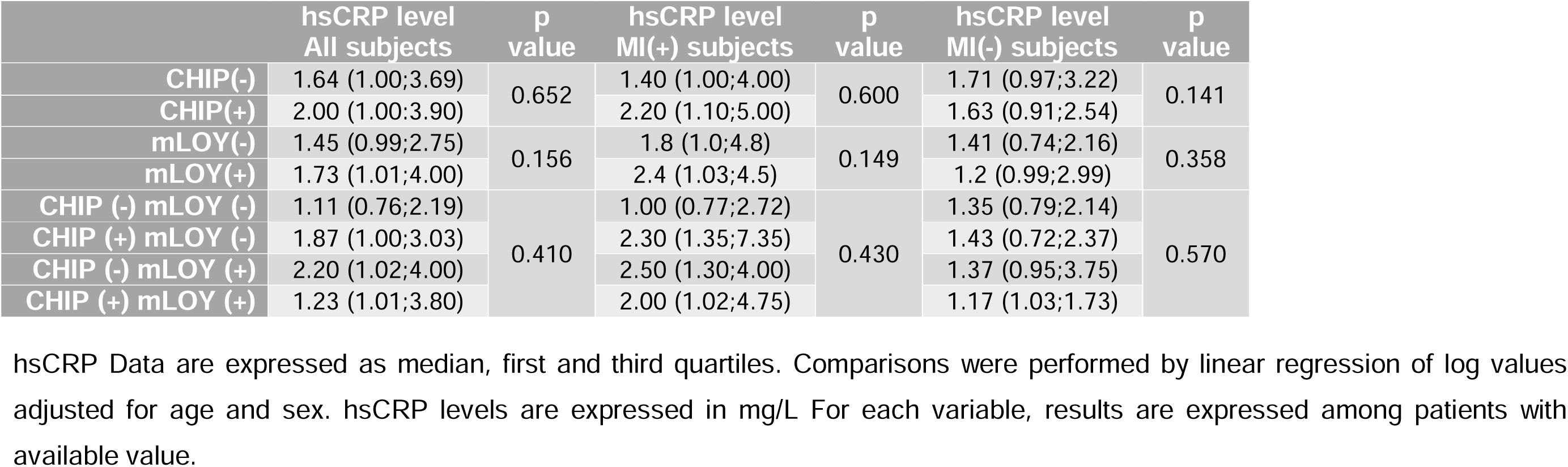
CHIP and mLOY are not associated with increased hsCRP level.

|  | hsCRP level<br>All subjects | p<br>value | hsCRP level<br>MI(+) subjects | p<br>value | hsCRP level<br>MI(-) subjects | p<br>value |
| --- | --- | --- | --- | --- | --- | --- |
| CHIP(-) | 1.64 (1.00;3.69) | 0.652 | 1.40 (1.00;4.00) | 0.600 | 1.71 (0.97;3.22) | 0.141 |
| CHIP(+) | 2.00 (1.00;3.90) |  | 2.20 (1.10;5.00) |  | 1.63 (0.91;2.54) |  |
| mLOY(-) | 1.45 (0.99;2.75) | 0.156 | 1.8 (1.0;4.8) | 0.149 | 1.41 (0.74;2.16) | 0.358 |
| mLOY(+) | 1.73 (1.01;4.00) |  | 2.4 (1.03;4.5) |  | 1.2 (0.99;2.99) |  |
| CHIP (-) mLOY (-) | 1.11 (0.76;2.19) | 0.410 | 1.00 (0.77;2.72) | 0.430 | 1.35 (0.79;2.14) | 0.570 |
| CHIP (+) mLOY (-) | 1.87 (1.00;3.03) |  | 2.30 (1.35;7.35) |  | 1.43 (0.72;2.37) |  |
| CHIP (-) mLOY (+) | 2.20 (1.02;4.00) |  | 2.50 (1.30;4.00) |  | 1.37 (0.95;3.75) |  |
| CHIP (+) mLOY (+) | 1.23 (1.01;3.80) |  | 2.00 (1.02;4.75) |  | 1.17 (1.03;1.73) |  |

hsCRP Data are expressed as median, first and third quartiles. Comparisons were performed by linear regression of log values adjusted for age and sex. hsCRP levels are expressed in mg/L For each variable, results are expressed among patients with available value.

Very few data are available about atheroma burden associated with CHIP and/or mLOY in humans. Moreover, the modulation of the atherogenic effect of CHIP by mLOY remains unexplored. We thus asked whether CHIP alone or in association with mLOY was associated with an increased atherosclerotic burden. In MI(+) subjects we observed a similar proportion of multitroncular coronary lesions and carotid stenosis >50% in CHIP(+) and CHIP(-) subjects (Table 3). The global atheroma volume was explored by 3D ultrasound in 34 MI(+) patients, without difference between CHIP(+) and CHIP(-) subjects. Similar results were obtained when analyzing only CHIP(+) subjects carrying *TET2* and/or *DNMT3A* mutations (Supplementary Table S7), when comparing mLOY(+) and mLOY(-) subjects (Table 3), or when analyzing the association of CHIP with mLOY (Supplementary Table S9). Concordantly, in MI(-) subjects, all available atherosclerosis markers were similar between CHIP(+) and CHIP(-), as well as between mLOY(+) and mLOY(-) subjects (Table 3). This was also true when analyzing the effect of the different combinations of CHIP and mLOY (Supplementary Table S9). Once again, all these results were confirmed in subjects with a VAF ≥5% for CHIP or a mLOY≥50% (Supplementary Table S8), or when adjusting further on CVRF.

**Table 3:** CHIP and mLOY are not associated with an increased atherosclerotic burden.

| Atherosclerosis burden evaluation in MI(+) subjects |  |  |  |  |  |  |  |
| --- | --- | --- | --- | --- | --- | --- | --- |
|  | All patients<br>(n = 149) | CHIP (-)<br>(n = 70) | CHIP (+)<br>(n = 79) | p-value | mLOY (-)<br>(n = 53) | mLOY (+)<br>(n = 44) | p-value |
| Multitroncular lesions, n (%) | 68 (45.6) | 29 (41.4) | 39 (49.4) | 0.484 | 25 (47.2) | 20 (45.4) | 0.717 |
| Carotid stenosis ≥ 50%, n (%) | 7 (4.7) | 2 (2.8) | 5 (6.3) | 0.317 | 2 (3.8) | 3 (6.8) | 0.451 |
| Global atheroma volume<br>(mm<sup>3</sup>), median (Q1;Q3) | 499.5<br>(408.0;604.5) | 455.0<br>(374.0;555.0) | 520.0<br>(411.5;611.5) | 0.333 | 601.0<br>(412.0;718.0) | 492.0<br>(344.5;600.5) | 0.707 |
| Atherosclerosis burden evaluation in MI(-) subjects |  |  |  |  |  |  |  |
|  | All patients<br>(n = 297) | CHIP (-)<br>(n = 175) | CHIP (+)<br>(n = 122) | p-value | mLOY (-)<br>(n = 84) | mLOY (+)<br>(n = 39) | p-value |
| Patients with atherosclerotic<br>plaque, n (%) | 135 (45.4) | 81 (46.3) | 54 (44.3) | 0.997 | 34 (40.5) | 19 (48.7) | 0.537 |
| Number of plaque, median<br>(Q1;Q3) | 1 (1;2) | 2 (1;2) | 1 (1;2) | 0.258 | 2 (1;2) | 2 (1;2) | 0.863 |
| Intima Media Thickness (mm),<br>median (Q1;Q3) | 0.68<br>(0.60;0.76) | 0.67<br>(0.60;0.76) | 0.68<br>(0.59;0.74) | 0.897 | 0.67<br>(0.62;0.76) | 0.72<br>(0.57;0.83) | 0.706 |

Data are expressed as numbers and frequency or median, first and third quartiles. For quantitative values, comparisons were made by linear regression of log values adjusted for age and sex. For qualitative parameters, comparisons were made by the fisher test and logistic regression. For each variable, results are expressed among patients with available value.

Altogether, our results suggest that in both the context of a recent MI and in healthy individuals, CHIP is not associated with a systemic inflammation or an increased atherosclerotic burden. Additionally, mLOY does not modulate inflammatory parameters or atherosclerosis, even in the presence of CHIP.

### Neither CHIP, mLOY, nor their combination highly impact the incidence of MI

To decipher whether mLOY could impact the atherothrombotic risk associated with CHIP, we analyzed the incidence of MI during the follow-up of MI(-) subjects. Seventy-nine subjects developed a MI after inclusion in the study with a median delay of 5.00 years. Subjects with MI during follow-up did not differ significantly from those without MI in terms of demography or cardiovascular risk (Supplementary Table S10). Contrary to other studies, we did not observe an association between CHIP and an increased incidence of MI (HR 1.033 [0.657;1.625] after adjustment on age, sex and CVRF). In comparison, hypercholesterolemia and smoking tended to associate with stronger risk of incident MI (HR = 1.474 [0.758;2.866] and 1.866 [0.943;3.690], respectively). Similarly, neither mLOY nor the association between CHIP and mLOY were associated with an increased incidence of MI (Figure 2A-B, Supplementary Table S11). Concordantly, we did not observe any difference in the prevalence of CHIP, mLOY or their association between MI(-) subjects who suffered from a MI during follow up and those who did not (Supplementary Table S10). This was also the case when restricting the analysis to CHIP with a VAF ≥5%, to subjects with a proportion of cells with mLOY ≥50%, to CHIP associated with *DNMT3A* or *TET2* mutations (Supplementary Figure S3A-S3B, Supplementary Table S10), or when making further adjustment on CVRF. These results suggest that the atherothrombotic risk associated with CHIP is moderate, and is not modulated by its association with mLOY. Moreover, neither CHIP, mLOY nor their combination were significantly associated with atherothrombotic recurrence (Supplementary Table S11).

**Figure 2:**
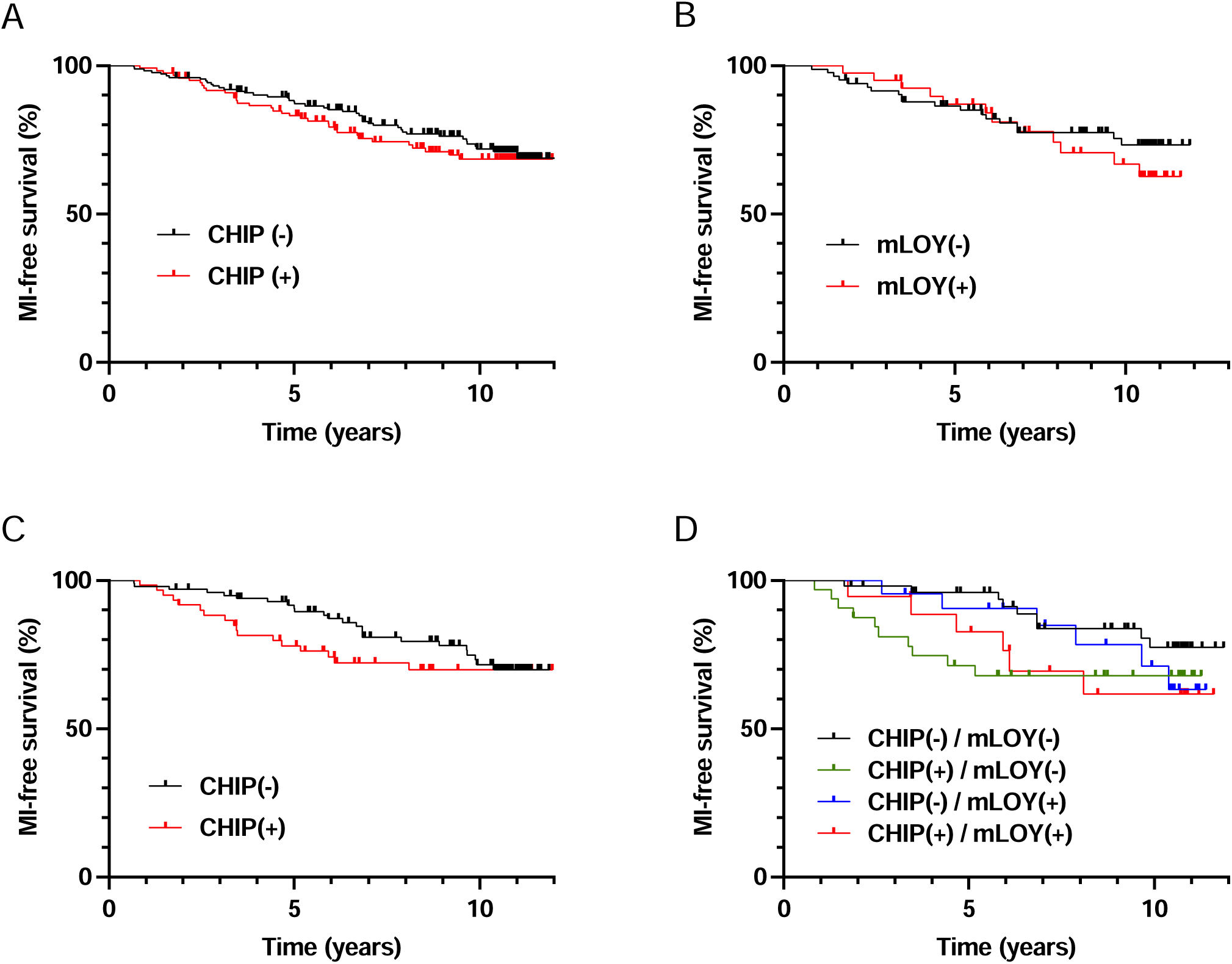
CHIP and mLOY do not increase significantly the risk of incident MI, but could accelerate it in male subjects. Incidence of MI during follow up according to the presence of CHIP (A) or mLOY (B) in MI(-) subjects. Incidence of MI during follow up according to the presence of CHIP (C) or the combination of CHIP and mLOY (D) in male MI(-) subjects. Survival was compared between the different groups with log-rank tests.

### CHIP in the absence of mLOY may accelerate the occurrence of MI

In order to search for a combined effect of CHIP with mLOY on the risk of incidence of MI, we finally focused our analysis on MI(-) male subjects. In this population, we observed that MI occurred earlier in CHIP(+) subjects, with a significant increased 5-year incidence of MI (log-rank test, p=0.014, Figure 2C). Such an effect was not observed in MI(-) female subjects (log-rank test, p=0.9402, Supplementary Figure S3C). Interestingly, this effect was more pronounced in CHIP(+)/mLOY(-) subjects who presented a significantly higher 5-year incidence of MI (log-rank test, p=0.010, Figure 2D) and a significantly lower median time to MI compared with other subjects (Kruskall-Wallis test, p=0.007). CHIP(+)/mLOY(-) subjects also presented a higher 5-year incidence of MI using Cox models after adjustment on age and CVRF (HR 7.81, p=0.0388). Altogether, our results suggest that CHIP do not increase the risk of MI, but may accelerate its incidence, particularly in the absence of mLOY.

## Discussion

Clonal hematopoiesis of indeterminate potential (CHIP) has previously been implicated in decreased overall survival, primarily due to its association with an increased incidence of cardiovascular diseases such as coronary artery disease (CAD) and stroke (Jaiswal et al., 2017, 2014). Experimental models have suggested that this association may arise from the pro-inflammatory phenotype of mutated monocytes/macrophages, contributing to the development of atherosclerotic plaques (Fuster et al., 2017; Jaiswal et al., 2017). However, recent findings in the literature have presented a complex and sometimes contradictory picture. Studies have reported varying results regarding the relationship between CHIP, inflammation markers, and atherothrombotic events, challenging the initial notion of a high atherothrombotic risk associated with CHIP (Bick et al., 2020; Busque et al., 2020; Kar et al., 2022; Kessler et al., 2022; Vlasschaert et al., 2023a). Moreover, there has been a scarcity of evidence linking CHIP to an increased burden of atherosclerosis in human subjects (Heimlich et al., 2024; Jaiswal et al., 2017; Wang et al., 2022; Zekavat et al., 2023). Additionally, limited data are available on the potential impact of chromosomal abnormalities, particularly mosaic loss of the Y chromosome (mLOY), on atherothrombosis in individuals with CHIP. In this study, we sought to investigate whether mLOY could modulate the effects of CHIP concerning systemic inflammation, atherosclerotic burden, and the risk of atherothrombotic events. To achieve this, we employed sensitive techniques, including targeted high-throughput sequencing and digital PCR, to analyze samples from two cohorts of meticulously phenotyped subjects.

In contrast to many previous studies, we conducted an analysis involving two distinct cohorts. The "cases" were individuals recruited from the CHAth study within 2 to 7 months following their first myocardial infarction (MI) after the age of 75. We also established a "control cohort" comprising 297 subjects from the 3C cohort, none of whom had experienced CVE before inclusion. This allowed us to assess the effects of CHIP and mLOY on inflammation and atherosclerosis independently of pre-existing cardiovascular disease. Our analysis revealed a remarkably high frequency of CHIP, with an estimated prevalence of 45% among our 446 subjects, with a median age of 76.4 years. This prevalence exceeded initial reports of 15-20% determined by whole exome sequencing (WES) in individuals aged 70-80,(Genovese et al., 2014; Jaiswal et al., 2014) likely attributable to the enhanced sensitivity of our sequencing technique. Importantly, our estimated prevalence aligns with studies employing similarly sensitive high-throughput sequencing techniques.(Guermouche et al., 2020; Mas-Peiro et al., 2020; Pascual-Figal et al., 2021; van Zeventer et al., 2021) Furthermore, our approach allowed us to reliably detect mutations with a variant allele frequency (VAF) as low as 1%. We chose to define subjects with detectable mutations at a VAF of 1% as CHIP(-), aligning with the WHO definition of CHIP.(Khoury et al., 2022) Notably, all our analyses also yielded identical results when comparing CHIP(+) patients to subjects without any detectable mutations or when comparing all patients with somatic mutations with a VAF≥1% to those without.

A recent study by Mas-Peiro et al. demonstrated an association between mLOY and increased post-transcatheter aortic valve replacement (TAVR) mortality (Mas-Peiro et al., 2023). In their investigation, a mLOY ratio threshold of 17% was deemed the most relevant for discriminating patients’ mortality risk. In our study, we employed a mLOY threshold of 9% to identify mLOY, a value determined to allow a reliable mLOY detection and to distinguish it from the background noise signal. Using this threshold, we frequently detected mLOY in male subjects within our cohort, with an estimated prevalence of 37.7% among 220 subjects. This prevalence surpasses that reported by Forsberg et al (Forsberg et al., 2014) but aligns with recent findings from studies employing sensitive techniques.(Zink et al., 2017)

Surprisingly, our cohort did not reveal any significant association between the presence of CHIP and the detection of mLOY. This contrasts with the results of two recent studies,(Ljungström et al., 2022; Zink et al., 2017) possibly explained by the heightened sensitivity of our methodology in reliably detecting both somatic mutations and mLOY, which may exist in very small proportions of blood cells.

Mouse models of CHIP have recently suggested that somatic mutations are linked to a pro-inflammatory phenotype of mutated monocytes/macrophages, an observation supported by human samples using single-cell RNA sequencing (Abplanalp et al., 2021). However, contradictory results have emerged when examining plasma markers of inflammation. Studies, including ours, have not consistently identified a significant increase in plasma hsCRP or proinflammatory cytokines in CHIP carriers (Cook et al., 2019; Pascual-Figal et al., 2021), whereas others have reported such associations (Bick et al., 2020; Busque et al., 2020). Importantly, our study did not show any significant association between the detection of CHIP(+/-)mLOY and plasma levels of IL1ß and IL6 in MI(+) subjects, suggesting that CHIP’s association with increased systemic inflammation may depend on specific stimulating factors. Notably, recent studies have reported elevated levels of plasma inflammatory markers in CHIP carriers during atherothrombotic events, such as MI or stroke (Arends et al., 2023; Böhme et al., 2022; Wang et al., 2022), indicating that CHIP may amplify systemic inflammation under specific conditions, but not necessarily in a basal state, as in our study.

To date, only a limited number of studies have successfully linked the presence of CHIP to an increased atherosclerotic burden. While Jaiswal *et al*. reported a higher calcic score in subjects with CHIP (Jaiswal et al., 2017), and Zekavat *et al*. suggested an increased atherosclerosis (Zekavat et al., 2023), these evaluations relied on self-reported atheroma and indirect parameters. In the context of coronary artery disease, only 2 studies addressed this question with conflicting results. Heimlich *et al* observed an increased prevalence of stenosis and obstructive stenosis in CHIP(+) subjects, particularly of the left main artery, while Wang *et al* did not notice any association between CHIP and the extent of coronary artery disease. (Heimlich et al., 2024; Wang et al., 2022) Our study stands as one of the first to utilize direct markers of atherosclerosis, including global atheroma volume, in CHIP(+) subjects within the context of coronary artery disease. Strikingly, we did not detect a clear increase in atherosclerosis among CHIP or mLOY carriers, either individually or in combination. Conversely, increased atherosclerotic burden associated with CHIP has been observed in patients with stroke.(Mayerhofer et al., 2023)

Our study bears some limitations, the first of them being a relatively modest sample size of 449 subjects, which did not allow us to establish a direct association between CHIP, either alone or in conjunction with mLOY, and coronary heart disease. These results are in contradiction with previous studies based on cohorts composed of a high number of subjects (Jaiswal et al., 2014, 2017; Vlasschaert et al., 2023a). At the time of our study’s initiation, the literature suggested a CHIP prevalence of 20% after 75 years and an increased MI risk associated with CHIP, with a hazard ratio of 1.7. Based on this data, a cohort of 112 cases would have been sufficient to demonstrate a more frequent presence of CHIP in MI(+) patients compared to MI(-) subjects with a power of 0.90 (more details are available in the Supplementary Methods). Although our study was not designed to demonstrate an association between CHIP and incident MI, we were able to confirm that the increased risk of MI associated with the presence of CHIP, if any, is lower 1.7, which is in accordance with more recent studies (Vlasschaert et al., 2023a; Zekavat et al., 2023; Zhao et al., 2024).

Different parameters could have also contributed to the discrepancy of our results with those of previous studies. First, the age of our subjects (≥75 years in the CHAth study, ≥65 years in the 3C study) is higher than the one of other cohorts. Jaiswal et al., 2017; Vlasschaert et al., 2023a; Zhao et al., 2024). Then our strategy to search for somatic variants was also different. In particular, we did not use the criteria defined by Vasschaert *et al* (Vlasschaert et al., 2023b) to cure variants that were called. This had a limited effect since 86.8% of the variants detected in our cohort were concordant with the criteria of Vlasschaert *et al* (Vlasschaert et al., 2023b), impacting the conclusion on the existence of a CHIP in only 15 patients. We also searched for a specific effect of *TET2* mutations on inflammation, atherosclerosis or risk of MI, as the literature suggests that they could be considered as “positive controls”. However, we did not find any significant effect of *TET2* mutations. Finally, we were not able to reliably detect variants with a VAF<1% and could have missed the effect of low-VAF variants, as recently shown by Zhao *et al* (Zhao et al., 2024).

However, our results align with previous studies that reported either no difference in CHIP prevalence between individuals with MI and those without (Busque et al., 2020), or no significant association between CHIP and incident *de novo* or recurrent atherothrombotic events (Arends et al., 2023; David et al., 2022; Kar et al., 2022). Recently, Kessler et al. proposed that the increased risk of atherothrombotic events might be limited to CHIP with a VAF of 10% or higher (Kessler et al., 2022). However, even when considering this criterion, the association was not validated in a cohort of 173,585 subjects. Moreover, in our cohort the observed effect of CHIP on the risk of MI (HR = 1.033) was substantially lower than the ones observed for other established cardiovascular risk factors such as hypercholesterolemia (HR = 1.475) or smoking (HR = 1.865). This underscores the formidable challenges of identifying associations with low effect sizes, necessitating cohorts comprising hundreds of thousands of subjects to achieve statistical significance. Collectively, these findings suggest that any atherothrombotic risk associated with CHIP is limited in scope and cannot be used in clinical practice for the management of patients with a history of cardiovascular disease or a risk of atherothrombosis.

However, we believe that our study has also some strength. Notably, we employed highly sensitive techniques for the reliable detection of both CHIP and mLOY, surpassing the capabilities of large cohort studies relying on whole exome sequencing and SNP arrays. Furthermore, we meticulously assessed atherosclerotic burden using sensitive parameters and evaluated two cohorts—one comprising MI(+) subjects and the other MI(-) subjects— with precise cardiovascular phenotyping at inclusion and rigorous follow-up, including direct evaluation and adjudication of all CVE.

In summary, our findings provide a nuanced perspective on the relationship between CHIP, mLOY, and cardiovascular outcomes, highlighting the need for larger, more comprehensive studies to elucidate potential associations further. Our findings that CHIP might expedite the onset of MI, particularly in the absence of mLOY warrant further investigation in larger subject cohorts.

## Data Availability

All data produced in the present work are contained in the manuscript.

## Acknowledgements

The authors would like to thank the University Hospital of Bordeaux CRB-K for processing the samples and Coralie Foucault and the Agilent Society for their partnership in supplying NGS reactive. We also thank Marina Migeon, Joël Decombe and Candice Falourd for their technical help, as well as Christel Duprat and Matthieu Meilhan for operational organization of the project.

## Authorship

TC, OM and CJ designed the study. SF, YP, TC, AG and JB enrolled the subjects in the CHAth study and performed all cardiovascular evaluation. AS, CT and SD set up the 3 City Study Cohort. AB, SM, OM analyzed the NGS data. MD, GM and DAT realized the statistical analysis. SF, SM, MD, DAT, CJ, OM and TC wrote the paper. All authors read and agreed to the final version of the manuscript.

## Author approval

All authors have seen and approved the manuscript.

## Registration

URL: https://clinicaltrials.gov; Unique identifier: NCT04581057.

