## Supplementary Methods for "Evaluation of Clonal Hematopoiesis and Mosaic Loss of Y Chromosome in Cardiovascular Risk: an analysis in prospective studies"

**Objectives and methodology to determine the number of subjects to include in the CHAth study**

The CHAth study was an observational transversal monocentric study. It aimed at evaluating the prevalence of CHIP in patients over 75 presenting with a first cardio-vascular event (CVE) and to determine if CHIPs are more frequent in this population compared to a control cohort without CVE (recruited from the Three-City study cohort or 3C).

To determine the number of patients necessary to achieve our objectives, we considered a CHIP prevalence of 20% in the general population after the age of 75 years, as estimated by Genovese et al (Genovese et al., 2014), and Jaiswal et al (Jaiswal et al., 2017, 2014). At this time the relative risk of MI associated with CHIP was shown to be 1.7, leading to an expected prevalence of CHIP of 37% in subjects who presented a MI. Based on these hypotheses, the recruitment of 112 patients in the CHAth was estimated to be sufficient to show a higher prevalence of CHIP in MI(+) patients compared to MI(-) subjects with a statistical power of 0.90 at a type I error rate of 5%.

Our study was not designed to show an effect of CHIP on incident MI during follow up, including in the 297 MI(-) subjects from the 3C study who were used as control subjects for MI(+) patients.

**Patients’ inclusion the CHAth study**

Eligible patients were $\geq$ 75 years of age and admitted for their first acute coronary event, without any history of other previous cardiovascular events. According to guidelines, a myocardial infarction was defined as a myocardial injury with clinical evidence of myocardial ischemia, with significant change in cTroponin levels above the 99^th^ percentile and at least one of the following criteria: symptoms of myocardial ischemia, ischemic ECG changes, development of pathological Q waves, imaging evidence of loss of viable myocardium or new regional wall motion abnormality, identification of a coronary thrombus by angiography.(Thygesen et al., 2019) Patients with type 1 myocardial infarctions, and type 2 if there was evidence of atherosclerotic coronary disease, were included. Those with myocardial infarctions due to vasospasm, microvascular dysfunction, non-atherosclerotic coronary dissection, or oxygen supply/demand imbalance alone were excluded.

The other inclusion criterion was the absence of hematological malignancy, known or revealed by blood count at admission at the time of the event. If an abnormality was detected in the blood count at the time of admission, a complementary analysis was performed to search for a non-neoplastic cause (hemoconcentration, dilution, inflammation due to an active and reversible condition, vitamin or iron deficiency, bleeding not related to an active neoplasia). If no reversible cause was found and/or if blood count was not normalized during the hospital stay, the patient was excluded from the study. Hemoglobin was considered normal if between 12 g/dL and 16 g/dL in women, and 13 g/dL and 17 g/dL in men; platelets count had to be between 150 and 400 G/L, and white blood cells were expected to be between 4 and 10 G/L.

Exclusion criteria encompassed the absence of atherosclerotic coronary disease, uncontrolled diabetes (defined as HbA1c > 10%), a previous atherosclerotic cardiovascular event before age 75 (myocardial infarction, ischemic stroke, peripheral arterial disease, angina pectoris), an hematological malignancy (known or revealed by blood count at admission), known chronic inflammatory disease (rheumatoid arthritis, arteritis, gastro-intestinal illness), infection or fever >38.5°C within 15 days prior to hospitalization, surgical operation within 30 days prior to hospitalization, the presence of an active neoplasia, and long-term use of anti-inflammatory therapies. Patients who were under legal protective measures, deprived of liberty by court order, unable to give their written informed consent, or having participated to an interventional study on a drug within 30 days before enrolment were also ineligible for enrollment in this study.

**Clinical and biological data reported in MI(+) and MI(-) subjects**

Data related to patient’s characteristics included: age, gender, body mass index. For traditional alterable cardiovascular risk factors, smoking was defined as having an active smoking habit or having stopped within the last three years. Hypertension was defined as a systolic blood pressure of at least 140 mmHg and/or a diastolic blood pressure of at least 90 mmHg from the means of daily measures during the hospital stay, or by the use of anti-hypertensive drugs. Dyslipidemia was defined as a c-LDL > 4.1 mmol/L (> 1.6 g/L) and/or c-HDL < 1.03 mmol/L (< 0.4 g/L) in men and < 1.29 mmol/L (< 0.5 g/L) in women, or by the use of lipid-lowering drugs.(Arnett et al., 2019) A threshold of HbA1c > 6.5% was used to define diabetes, as well as use of anti-diabetic agents.(Association, 2019)

For MI(+) subjects, following data were collected regarding the atherothrombotic event: existence of a ST-elevation; concerned territory; number of main vessels with a stenosis > 50%, according to morphological assessment by the cardiologist performing the coronary angiography; acute phase treatment (medical treatment, fibrinolysis, angioplasty, coronary artery bypass graft surgery); and presence and type of potential complications, including: heart failure (defined according to the 2016 ESC guidelines(Ponikowski et al., 2016)), ventricular arrythmia (defined as sustained ventricular tachycardia or ventricular fibrillation), high-degree atrio-ventricular block, pericarditis, mechanical complication comprising mitral regurgitation, ventricular free-wall rupture and ventricular septal defect, as well as intra-cardiac thrombosis diagnosed by echocardiography. At inclusion, a routine cardiovascular evaluation was performed at inclusion (between 2 and 7 month after MI occurrence). Subjects were asked about presence of dyspnea or angina, evaluated with NYHA and CCS scales. Information was obtained from medical records about a potential recurrence of a cardiovascular event since the index event (new MI, coronary revascularization, stroke, hospitalization for acute heart failure). Traditional CVRF were noted. Routine biological analyses comprising blood count, high-sensitive CRP (hsCRP), lipid profile, HbA1c were performed in the laboratory of the university hospital of Bordeaux on fresh samples. A blood sample (EDTA) was drawn and sent within 4 hours to the Biological Cancer Resources Center of the University Hospital of Bordeaux.

For MI(-) subjects, data available at inclusion included hsCRP level, creatinine, lipid profile and traditional CVRF. No blood count was available but none of the subjects developed cancer (including hematological malignancy) during follow-up suggesting that detectable somatic mutations were indicative of CHIP and not hematological malignancy.

**Follow up of subjects**

For MI(+) subjects, the follow-up was conducted with a standardized questionnaire previously validated in clinical trials.(Lafitte et al., 2013) Recurrence of MI as well as any significant cardiovascular event (cardiovascular death, acute coronary syndrome, stroke or transient ischemic attack, congestive heart failure, secondary coronary revascularization, or peripheral vascular surgery) occurring between the initial event and the year after inclusion in the study were recorded. All medical records of participants who died, or who reported on the questionnaire that they had experienced cardiovascular symptoms between baseline and follow-up evaluations, were reviewed by one of the investigators, and the patient practitioners were contacted. For MI(-) subjects, a follow up was performed for up to 12 years. All cardiovascular events were recorded (including MI) with adjudication upon medical records by an expert committee (Figure 1).

**NGS sequencing and analysis**

*Library preparation and sequencing*

A custom RNA-baits panel was designed to cover 56 genes involved in myeloid malignancies. The list of the genomic regions targeted is available in the Supplementary Table 1. Libraries were prepared from 200 ng of DNA for each patient using SureSelect XT Low Input kit (Agilent) or using Magnis NGS Prep System (Agilent). Libraries were pooled for multiplex sequencing on a NextSeq500 (Illumina) with Mid Output Kit v2.

*Bioinformatic pipeline*

We developed a bioinformatic pipeline to analyze sequencing data in order to control each step of analysis. FASTQ files were generated by bcl2fastq. They were then aligned against reference genome hg19 (2013) with bwa-mem, producing BAM files. After this step, duplicate reads were tagged but not removed using agilent locatit software. Finally, coverage analysis was performed with bbctools, mosdepth, samtools and resulting metrics were gathered with MultiQC to assess sequencing data quality, including depth of coverage for every gene in the panel. For this study we used 3 different tools for variant calling in order to detect with good accuracy mutations in samples. We have chosen GATK Mutect2, VarScan and VarDict to detect somatic mutations. Annovar software (version 2020-06-08) and ensembl VEP (v.103) were used to annotate all called variants. The following databases were used:

- COSMIC 92 (Catalogue Of Somatic Mutations In Cancer)

- gnomAD 2.1.1 (The Genome Aggregation Database), ExaC (The Exome Aggregation Consortium), 1000 genome, ESP (Exome Sequencing Project) to assess variant frequency in world population

- SIFT, PolyPhen2, PROVEAN (in-silico pathology prediction tools)

- dbSNP (The Single Nucleotide Polymorphism database)

- ClinVar (2021-01-03) (information on the relationships between variants and human health): this descriptor provides information on the clinical effect of the variant

- InterVar (software tool for automatic clinical interpretation of genetic variants by the ACMG/AMP 2015 guideline).

We observed a median depth of sequencing of 2111X [1578;2574] for all regions sequenced. For the “prototypical” CHIP genes defined by Vlasschaert *et al* (Vlasschaert et al., 2023), the sequencing depth was 2694X [1875;3785] for patients from the CHAth study and 3455X [2266;4885] for patients from the 3C study. More specifically, for *DNMT3A* and *TET2* genes, the median depths of sequencing were 2531X [1818 ;3313] and 3710X [2444 ;4901] for patients from the CHAth and 3C studies respectively.

*Review and classification of mutations*

Intronic and synonymous mutations were removed as well as variants with VAF <1% and variants with a minor allele frequency (MAF) ≥0.1% listed in databases (dbSNP, gnomAD, ExaC, 1000 genome, ESP). Variants known as recurring artifact (manually curated and stocked in a local database) were also removed. Finally, retained variants were reviewed independently by 2 molecular biologists for (i) visual inspection of reads in BAM file to determine if they were real mutations or artifacts and for (ii) classification of the pathogenicity of mutations. To be interpretable, the generated data had to meet the following criteria:

- Minimum read depth of 200X

- Sufficient coverage of the different regions in both reading directions

- Total number of reads corresponding to the variant must be greater than 10.

In order to have more confidence in variants with low VAF, we distinguished them from artifacts by estimating the background noise *via* the median absolute deviation and the corresponding confidence intervals. To exclude variants that could represent artifacts, we only retained those that presented a VAF at least 2-fold higher than the upper limit of the confidence interval.

The classification of mutations was based on the consensus recommendation of the Association for Molecular Pathology and the American Society of Clinical Oncology.(Li et al., 2017) Variants were classified as pathogenic, likely pathogenic or of unknown significance according to the criteria shown in Supplemental Table 2.

Although our NGS panel and criteria for the classification of mutations are classical for the study of hematological malignancies, it should be noticed that they do not strictly corresponding to the criteria defined by Vlasschaert *et al* (Vlasschaert et al., 2023) who recently defined a list of genes/mutations considered to be prototypical CHIP-genes more tightly associated with cardiovascular disease.

**Detection of mLOY by digital droplets PCR**

The search of mLOY was performed thanks to an in-house droplet digital PCR technic using the following primers and probes:

- Primer-amel-Fwd : 5’- CCCCTGGGCACTGTAAAGAAT
- Primer-amel-Rev: 5’- CCAAGCATCAGAGCTTAAACTG
- Probe-amelX : 5’- HEX-CCAAATAAAGTGGTTTCTCAAGT-BHQ
- Probe-amelY: 5’- FAM-CTTGAGAAACATCTGGGATAAAG-BHQ.

Briefly, 75 ng of DNA was mixed with ddPCR supermix for Probes (no dUTP, Biorad), primers (0.9µM each) and probes (0.25µM each). The emulsion was prepared with the QX-100 (Biorad). The amplification program was as follows: 10-minutes denaturation at 95°C, followed by 40 cycles of 30 seconds at 94°C, 1 minute at 55°C, and inactivation of 10 minutes at 98°C. The number of droplets positive for amelX and amelY was determined on the QX-200 droplet reader (Biorad) using the Quantasoft software version 1.5 (Biorad). At least 10,000 droplets were analyzed in each well. The results were expressed as the estimated percentage of cells carrying a mLOY based on the copy number of amelX and amelY genes. We validated our technic by studying 30 men of less than 40 years who had a normal karyotype as assessed by conventional cytogenetic study. We determined a cut off of >9% of cells with mLOY as indicative the presence of a mLOY. We considered the level of mLOY as “low” when the proportion of cells with a mLOY was between 9 and 50%, and a “high” when the proportion of cells with mLOY was above 50%.

Thygesen K, Alpert JS, Jaffe AS, Chaitman BR, Bax JJ, Morrow DA, White HD, ESC Scientific Document Group, Thygesen K, Alpert JS, Jaffe AS, Chaitman BR, Bax JJ, Morrow DA, White HD, Mickley H, Crea F, Van de Werf F, Bucciarelli-Ducci C, Katus HA, Pinto FJ, Antman EM, Hamm CW, De Caterina R, Januzzi JL, Apple FS, Alonso Garcia MA, Underwood SR, Canty JM, Lyon AR, Devereaux PJ, Zamorano JL, Lindahl B, Weintraub WS, Newby LK, Virmani R, Vranckx P, Cutlip D, Gibbons RJ, Smith SC, Atar D, Luepker RV, Robertson RM, Bonow RO, Steg PG, O’Gara PT, Fox KAA, Hasdai D, Aboyans V, Achenbach S, Agewall S, Alexander T, Avezum A, Barbato E, Bassand J-P, Bates E, Bittl JA, Breithardt G, Bueno H, Bugiardini R, Cohen MG, Dangas G, de Lemos JA, Delgado V, Filippatos G, Fry E, Granger CB, Halvorsen S, Hlatky MA, Ibanez B, James S, Kastrati A, Leclercq C, Mahaffey KW, Mehta L, Müller C, Patrono C, Piepoli MF, Piñeiro D, Roffi M, Rubboli A, Sharma S, Simpson IA, Tendera M, Valgimigli M, van der Wal AC, Windecker S, Chettibi M, Hayrapetyan H, Roithinger FX, Aliyev F, Sujayeva V, Claeys MJ, Smajić E, Kala P, Iversen KK, El Hefny E, Marandi T, Porela P, Antov S, Gilard M, Blankenberg S, Davlouros P, Gudnason T, Alcalai R, Colivicchi F, Elezi S, Baitova G, Zakke I, Gustiene O, Beissel J, Dingli P, Grosu A, Damman P, Juliebø V, Legutko J, Morais J, Tatu-Chitoiu G, Yakovlev A, Zavatta M, Nedeljkovic M, Radsel P, Sionis A, Jemberg T, Müller C, Abid L, Abaci A, Parkhomenko A, Corbett S. 2019. Fourth universal definition of myocardial infarction (2018). *Eur Heart J* **40**:237–269. doi:10.1093/eurheartj/ehy462

Vlasschaert C, Mack T, Heimlich JB, Niroula A, Uddin MM, Weinstock J, Sharber B, Silver AJ, Xu Y, Savona M, Gibson C, Lanktree MB, Rauh MJ, Ebert BL, Natarajan P, Jaiswal S, Bick AG. 2023. A practical approach to curate clonal hematopoiesis of indeterminate potential in human genetic data sets. *Blood* **141**:2214–2223. doi:10.1182/blood.2022018825
