## Supplementary Figures for "Evaluation of Clonal Hematopoiesis and Mosaic Loss of Y Chromosome in Cardiovascular Risk: an analysis in prospective studies"

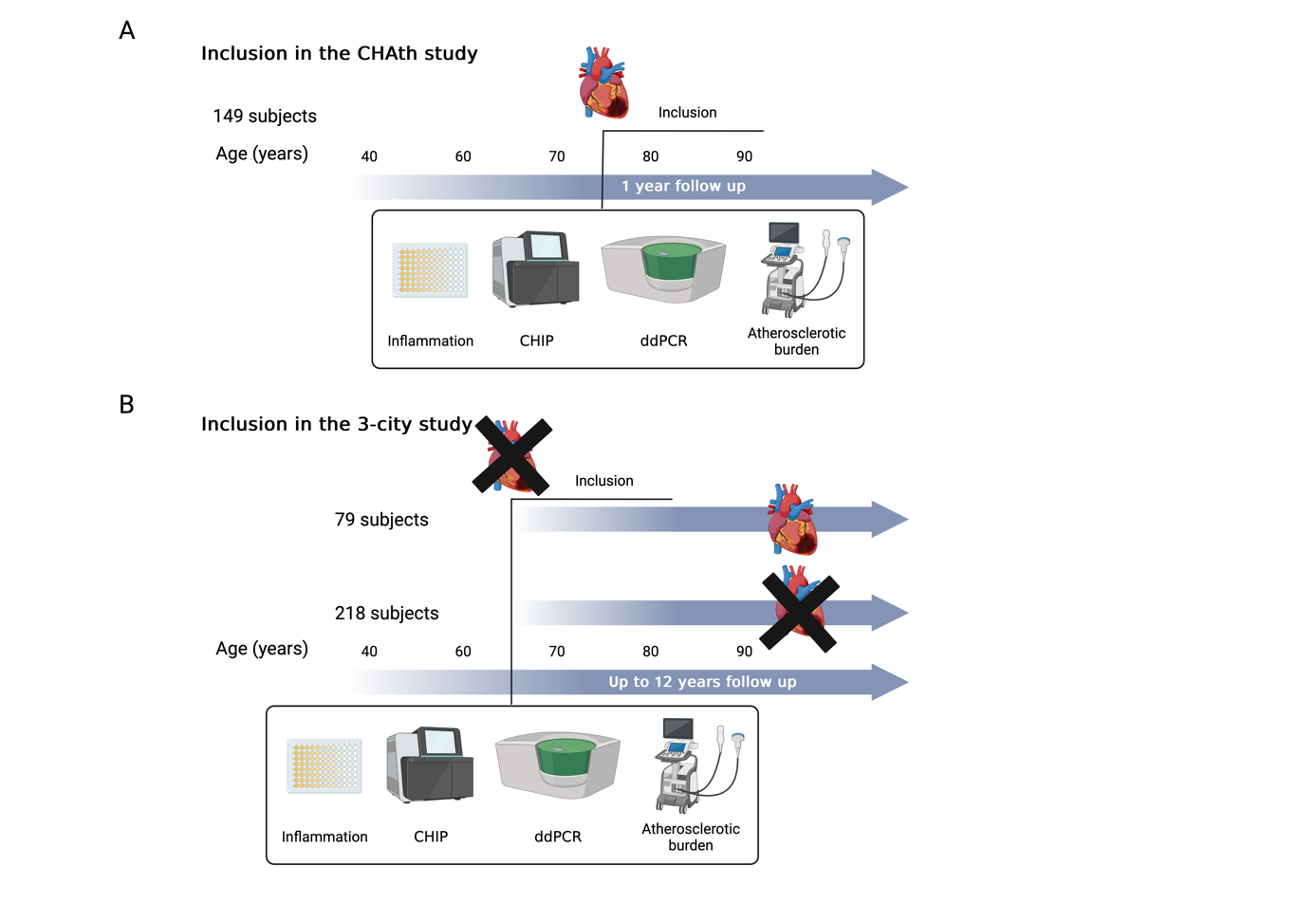

**Supplementary Figure S1: Sequence of testing realized in subjects included in the CHAth and 3C studies**

A: Subjects included in the CHAth study were enrolled after a first MI occurring after 75 years. Inflammatory parameters, atherosclerosis burden and detection of CHIP and mLOY were performed on samples obtained 2 to 7 months after MI. These subjects benefitted from a follow up of 1 year.

B: 297 subjects from the 3-city cohort were studied (inclusion at 65 years or more after selection upon electoral lists). None of them experienced CVE before inclusion. Inflammatory parameters, atherosclerosis burden and detection of CHIP and mLOY were performed on samples obtained at inclusion. These subjects benefitted from a follow up of up to 12 years. Seventy-nine suffered from a MI during follow up while 218 did not.

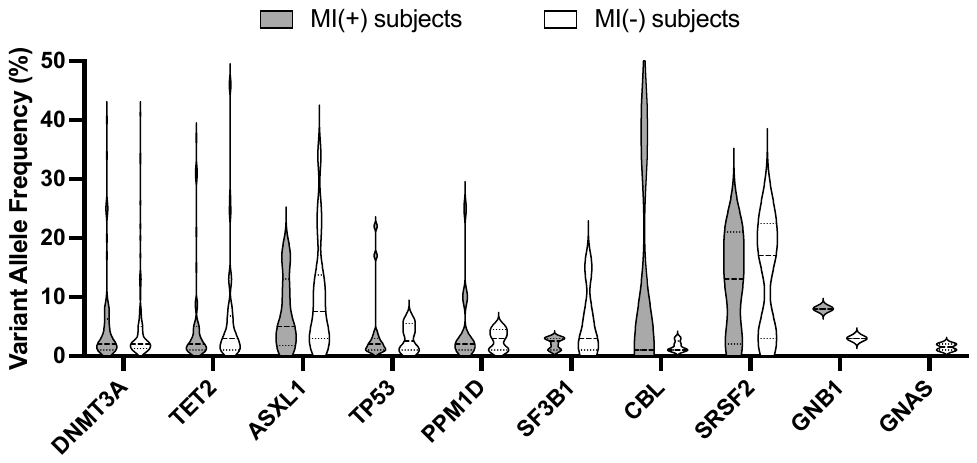

**Supplementary Figure S2: VAF of somatic mutations detected in MI(+) and +MI(-) subjects**

The graph represent the VAF measured for the different mutations detected in MI(+) and MI(**-**) subjects in the indicated genes

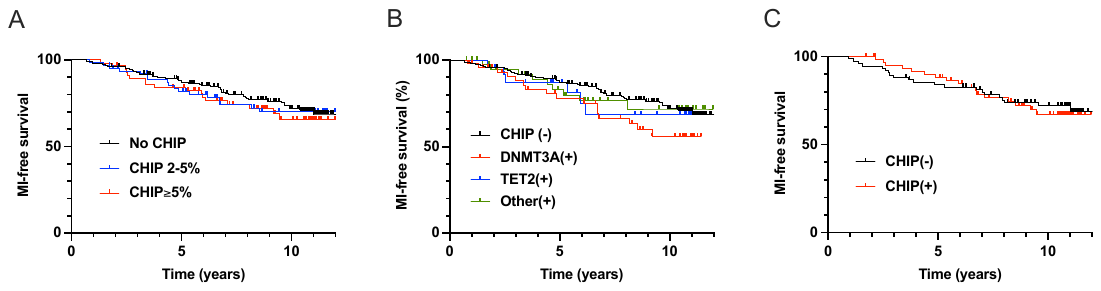

**Supplementary Figure S3: CHIP do not increase significantly the risk of incident MI**

Incidence of MI during follow up according to the VAF (A) or the mutated gene (B) in MI(-) subjects. Incidence of MI during follow up according to the presence of CHIP in MI(-) female subjects (C). Survival were compared between the different groups with log-rank tests.
