## Supplementary Tables for "Evaluation of Clonal Hematopoiesis and Mosaic Loss of Y Chromosome in Cardiovascular Risk: an analysis in prospective studies"

**Supplementary Table S1. Targeted sequencing panel used in the laboratory of Hematology of the University Hospital of Bordeaux for the diagnosis and follow-up of myeloid hematological malignancies and used in this study**

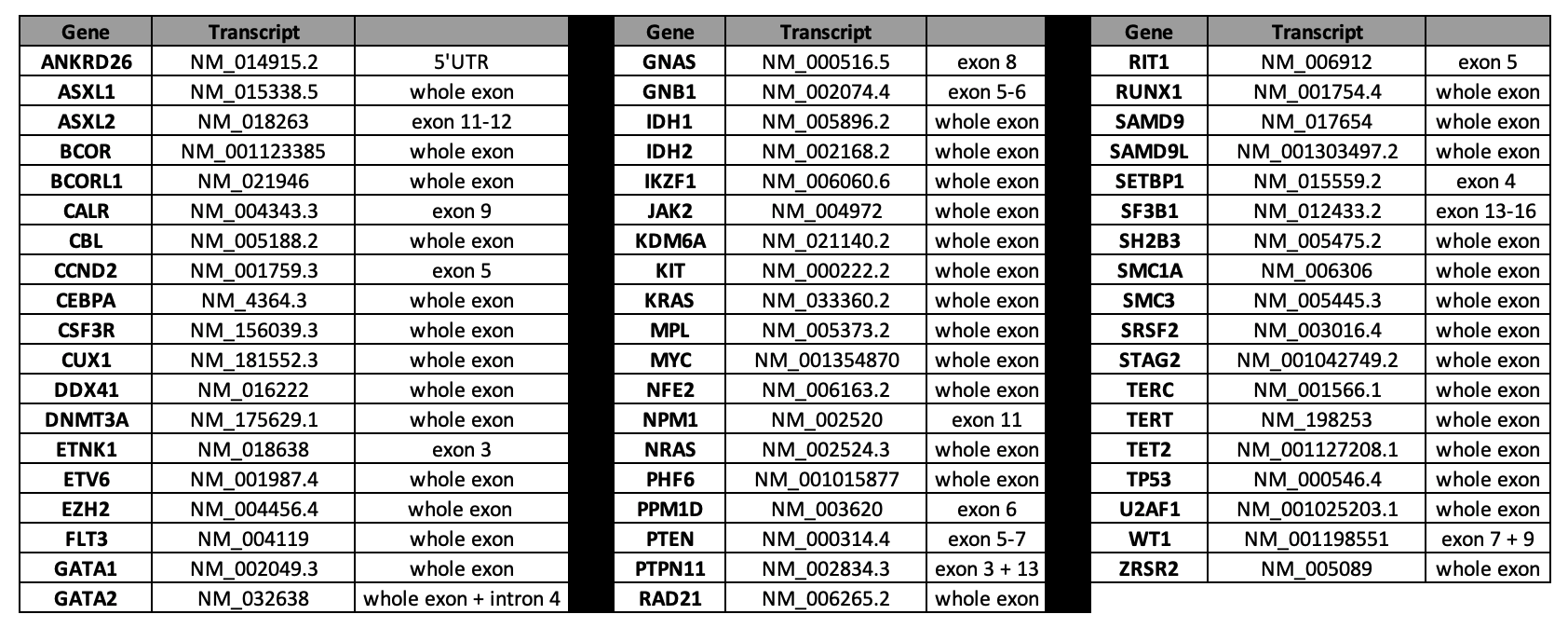

**Supplementary Table S2. Characteristics of the identified mutations according to the degree of certainty of their pathogenicity (deleterious, possibly deleterious or variant of undetermined significance)**

| Deleterious variant (A)* | - Variant resulting in a truncated protein  - Variant described in myeloid hematological malignancy (COSMIC) as somatic or with a VAF in favor of a somatic origin (<40% or between 60% and 80%)  - Variant with demonstrated functional effect (including splicing) |
| --- | --- |
| Possibly deleterious variant (B)* | - Variant probably not deleterious, with MAF <0.1% and VAF <40% or >60% (probably somatic origin) |
| Variant of undetermined significance (C) | - Variant probably not deleterious with VAF between 40 and 60% or equal to 100% (probably rare SNP)  - Splice variant predicted without impact by Human Slicing Finder |

Exceptions :

- *ASXL1*/*ASXL2/TET2:* only variants resulting in a truncated protein were retained as deleterious variant (A). For *TET2*, missense mutations were retained only when they affected CD1 or CD2 domain
- *ANKRD26*: non-coding region 5’-UTR was retained
- *TP53:* database IARC (https://p53.iarc.fr/) was used for the classification

* Only variants classified as A or B are retained as CHIP in the study.

**Supplementary Table S3. Mutational profile of CHIP(+) MI(+) subjects**

| **Patient** | **Gene** | **Transcript** | **c.** | **p.** | **Freq** | **Var.Cov.** | **Depth** | **COSMIC** | dbSNP |
| --- | --- | --- | --- | --- | --- | --- | --- | --- | --- |
| **002-GP** | TET2 | NM_001127208.2 | c.3621_3622dup | p.(Lys1208Argfs*19) | **31** | 499 | 1623 |  |  |
|  | TET2 | NM_001127208.2 | c.4355del | p.(Arg1452Glnfs*6) | **2** | 51 | 2539 |  |  |
| **003-CC** | DNMT3A | NM_022552.4 | c.2409-1G>C | p.? | **2** | 26 | 1362 |  |  |
|  | DNMT3A | NM_022552.4 | c.1136G>A | p.(Arg379His) | **6** | 48 | 823 | COSM5772441 |  |
|  | CBL | NM_005188.3 | c.1292T>C | p.(Val431Ala) | **1** | 29 | 2121 |  |  |
|  | ZRSR2 | NM_005089.3 | c.1041C>G | p.(Tyr347*) | **1** | 21 | 1888 |  |  |
|  | STAG2 | NM_001042749.2 | c.3023del | p.(Ser1008Leufs*19) | **1** | 16 | 1365 |  |  |
| **006-LE** | DNMT3A | NM_022552.4 | c.674del | p.(Asn225Metfs*91) | **2** | 16 | 867 |  |  |
| **008-ML** | TP53 | NM_001126112.2 | c.1182A>G | p.(*394Trpext*9) | **3** | 55 | 2118 |  |  |
| **009-JG** | SRSF2 | NM_003016.4 | c.284C>A | p.(Pro95His) | **2** | 29 | 1743 | COSM211029,COSM211504 | rs751713049 |
| **010-MG** | DNMT3A | NM_022552.4 | c.1752C>A | p.(Tyr584*) | **2** | 45 | 1810 | COSM5574186 |  |
|  | CBL | NM_005188.4 | c.2326G>A | p.(Val776Ile) | **1** | 36 | 3361 |  |  |
| **011-AD** | PPM1D | NM_003620.4 | c.1535del | p.(Asn512Ilefs*2) | **3** | 115 | 3888 | COSM267241 |  |
| **012-GW** | DNMT3A | NM_022552.4 | c.2357C>A | p.(Ser786*) | **8** | 85 | 1114 |  |  |
| **0015-RD** | DNMT3A | NM_022552.5 | c.2189_2197del | p.(Leu730_Glu733delinsGln) | **2** | 44 | 2238 |  |  |
| **0016-CA** | DNMT3A | NM_022552.5 | c.901C>T | p.(Arg301Trp) | **34** | 1131 | 3326 | COSM2911819 |  |
|  | TET2 | NM_001127208.3 | c.1001dup | p.(Pro335Serfs*5) | **8** | 462 | 5867 |  |  |
| **0017-GS** | DNMT3A | NM_022552.5 | c.2063G>A | p.(Arg688His) | **1** | 19 | 1328 | COSM4169953 | rs369713081 |
|  | DNMT3A | NM_022552.5 | c.1687G>A | p.(Val563Met) | **3** | 28 | 980 | COSM1583086 |  |
|  | DNMT3A | NM_022552.5 | c.1220T>C | p.(Ile407Thr) | **4** | 20 | 537 | COSM249801 | rs750108934 |
| **0018-CS** | TET2 | NM_001127208.3 | c.3804-1G>C | p.? | **1** | 34 | 2422 |  |  |
|  | TET2 | NM_001127208.3 | c.4537+1G>A | p.? | **3** | 65 | 2117 | COSM4170056 |  |
| **0019-MQ** | DNMT3A | NM_022552.5 | c.2458G>C | p.(Glu820Gln) | **20** | 340 | 1729 | COSM4768018 |  |
|  | TET2 | NM_001127208.3 | c.3752C>T | p.(Thr1251Met) | **1** | 39 | 3052 |  |  |
| **0020-LV** | DNMT3A | NM_022552.5 | c.2645G>A | p.(Arg882His) | **3** | 61 | 2282 | COSM52944 | rs147001633 |
|  | PPM1D | NM_003620.4 | c.1440dup | p.(Ala481Serfs*8) | **2** | 107 | 4567 |  |  |
| **021-MD** | TET2 | NM_001127208.3 | c.4138C>T | p.(His1380Tyr) | **2** | 58 | 2761 | COSM87161 | rs577810844 |
| **022-RR** | GATA2 | NM_032638.5 | c.1438G>C | p.(Gly480Arg) | **20** | 772 | 3834 |  |  |
|  | TP53 | NM_001126112.3 | c.944del | p.(Ser315Phefs*30) | **2** | 51 | 3106 |  |  |
|  | TP53 | NM_001126112.3 | c.707A>G | p.(Tyr236Cys) | **22** | 628 | 2894 | COSM10731 | rs730882026 |
|  | TP53 | NM_001126112.3 | c.626_627del | p.(Arg209Lysfs*6) | **1** | 58 | 4298 |  | rs1057517840 |
|  | TP53 | NM_001126112.3 | c.472_489del | p.(Arg158_Tyr163del) | **17** | 596 | 3593 |  |  |
| **023-MF** | DNMT3A | NM_022552.5 | c.2727del | p.(Phe909Leufs*13) | **2** | 47 | 2576 |  |  |
|  | DNMT3A | NM_022552.5 | c.1937-1G>C | p.? | **6** | 277 | 4497 |  |  |
| **024-CD** | CBL | NM_005188.4 | c.1259G>C | p.(Arg420Pro) | **1** | 16 | 1455 | COSM249797 |  |
| **025-PR** | TET2 | NM_001127208.3 | c.4567C>T | p.(Gln1523*) | **1** | 38 | 3060 | COSM96925 |  |
| **028-AG** | DNMT3A | NM_022552.5 | c.1103C>T | p.(Ala368Val) | **25** | 581 | 2288 |  | rs759087082 |
|  | PPM1D | NM_003620.4 | c.1636dup | p.(Leu546Profs*6) | **25** | 1086 | 4288 | COSM7346322 |  |
| **029-JL** | **DNMT3A** | **NM_022552.5** | c.2099C>T | p.(Pro700Leu) | 3 | 93 | 2938 |  | rs772368909 |
|  | **DNMT3A** | **NM_022552.5** | c.1232_1238delinsCGGGGGA | p.(Leu411_Gly413delinsProGlyAsp) | 4 | 50 | 1166 |  |  |
|  | **ZRSR2** | **NM_005089.4** | c.1361G>A | p.(Arg454Gln) | 4 | 45 | 1027 | COSM6970291 | rs746617360 |
| **031-JL** | DNMT3A | NM_022552.5 | c.2622T>A | p.(Tyr874*) | **1** | 20 | 1833 |  |  |
|  | DNMT3A | NM_022552.5 | c.2543T>C | p.(Phe848Ser) | **1** | 45 | 3169 |  |  |
|  | TET2 | NM_001127208.3 | c.162G>A | p.(Trp54*) | **2** | 60 | 2831 |  |  |
|  | TET2 | NM_001127208.3 | c.3347del | p.(Ile1116Lysfs*21) | **1** | 21 | 1826 |  |  |
| **032-JD** | SF3B1 | NM_012433.4 | c.1998G>C | p.(Lys666Asn) | **3** | 76 | 2586 | COSM132937 | rs377023736 |
|  | SF3B1 | NM_012433.4 | c.1997A>C | p.(Lys666Thr) | **1** | 44 | 2983 | COSM131556 | rs374250186 |
|  | SF3B1 | NM_012433.4 | c.1996A>C | p.(Lys666Gln) | **3** | 72 | 2608 | COSM132950 | rs754688962 |
|  | GATA2 | NM_032638.5 | c.1096G>T | p.(Gly366Trp) | **3** | 68 | 2473 |  |  |
|  | NFE2 | NM_006163.3 | c.194dup | p.(Tyr65*) | **4** | 102 | 2858 |  | rs774919590 |
|  | SH2B3 | NM_005475.3 | c.1141C>T | p.(Gln381*) | **1** | 12 | 1307 |  |  |
|  | TP53 | NM_001126112.3 | c.524G>A | p.(Arg175His) | **4** | 94 | 2660 | COSM10648 | rs28934578 |
|  | PPM1D | NM_003620.4 | c.1538del | p.(Leu513*) | **10** | 437 | 4368 |  |  |
| **034-MP** | TET2 | NM_001127208.3 | c.5641C>T | p.(His1881Tyr) | **3** | 98 | 3263 | COSM4383970 |  |
|  | SAMD9L | NM_001303497.3 | c.4086T>G | p.(Asn1362Lys) | **6** | 239 | 4074 |  |  |
| **035-JB** | DNMT3A | NM_022552.5 | c.2202C>G | p.(Phe734Leu) | **1** | 27 | 2376 |  |  |
| **036-LD** | DNMT3A | NM_022552.5 | c.2706del | p.(Phe902Leufs*4) | **2** | 40 | 2503 | COSM1166763 |  |
| **037-MD** | TET2 | NM_001127208.3 | c.5635G>C | p.(Glu1879Gln) | **5** | 167 | 3344 |  | rs944233155 |
| **038-JO** | TET2 | NM_001127208.3 | c.3804-2_3804-4delinsC | p.? | **32** | 594 | 1883 |  |  |
| **039-FL** | TET2 | NM_001127208.3 | c.872_873dup | p.(Val292Trpfs*2) | **1** | 38 | 3087 |  |  |
|  | BCOR | NM_001123385.2 | c.1271C>T | p.(Pro424Leu) | **22** | 693 | 3221 |  |  |
| **040-YF** | TET2 | NM_001127208.3 | c.5560T>G | p.(Phe1854Val) | **2** | 70 | 4538 |  |  |
|  | TP53 | NM_001126112.3 | c.658T>C | p.(Tyr220His) | **1** | 24 | 1621 | COSM44637 | rs530941076 |
|  | ASXL1 | NM_015338.6 | c.2893C>T | p.(Arg965*) | **1** | 74 | 5034 | COSM267971 | rs397515401 |
|  | BCORL1 | NM_021946.5 | c.4690G>A | p.(Val1564Met) | **2** | 49 | 2497 |  |  |
| **041-DB** | DNMT3A | NM_022552.5 | c.2314T>C | p.(Phe772Leu) | **25** | 593 | 2344 |  |  |
| **042-MJ** | DNMT3A | NM_022552.5 | c.2196T>G | p.(Phe732Leu) | **3** | 63 | 2162 | COSM5878764 |  |
| **043-PS** | ASXL1 | NM_015338.6 | c.1900_1922del | p.(Glu635Argfs*15) | **18** | 510 | 2907 | COSM36165 |  |
| **044-TF** | DNMT3A | NM_022552.5 | c.2173+1G>A | p.? | **2** | 43 | 2341 | COSM6907619 | rs763716866 |
|  | PPM1D | NM_003620.4 | c.1654C>T | p.(Arg552*) | **1** | 35 | 2907 | COSM982226 | rs779070661 |
| **048-CD** | DNMT3A | NM_022552.5 | c.2332G>A | p.(Val778Met) | **2** | 156 | 6556 |  |  |
|  | DNMT3A | NM_022552.5 | c.2195T>C | p.(Phe732Ser) | **7** | 543 | 8141 | COSM4383542 |  |
|  | DNMT3A | NM_022552.5 | c.1982T>G | p.(Ile661Ser) | **2** | 395 | 21402 |  |  |
| **049-MD** | GNB1 | NM_002074.5 | c.239T>C | p.(Ile80Thr) | **8** | 636 | 7733 | COSM1290046 | rs752746786 |
|  | ASXL1 | NM_015338.6 | c.1357G>T | p.(Glu453*) | **16** | 1947 | 12366 |  |  |
| **050-LB** | DNMT3A | NM_022552.5 | c.1978T>C | p.(Tyr660His) | **5** | 732 | 15101 | COSM6907810 |  |
| **055-BM** | DNMT3A | NM_022552.5 | c.2524_2525del | p.(Gln842Glyfs*12) | **24** | 726 | 3013 |  |  |
|  | SETBP1 | NM_015559.3 | c.2753G>A | p.(Arg918Gln) | **1** | 28 | 2690 |  | rs781033681 |
| **056-PD** | TET2 | NM_001127208.3 | c.3743T>C | p.(Leu1248Pro) | **37** | 984 | 2694 | COSM211718 | rs372179780 |
|  | TET2 | NM_001127208.3 | c.4546C>T | p.(Arg1516*) | **1** | 30 | 2312 | COSM43420 | rs370735654 |
|  | CBL | NM_005188.4 | c.1103A>G | p.(Tyr368Cys) | **38** | 385 | 1022 | COSM5749255 |  |
| **057-MN** | TP53 | NM_001126112.3 | c.712T>A | p.(Cys238Ser) | **2** | 62 | 3599 | COSM43700 |  |
| **058-JB** | TP53 | NM_001126112.3 | c.818G>A | p.(Arg273His) | **2** | 62 | 3322 | COSM10660 | rs28934576 |
|  | TP53 | NM_001126112.3 | c.817C>T | p.(Arg273Cys) | **1** | 49 | 4007 | COSM10659 | rs121913343 |
| **059-LB** | DNMT3A | NM_022552.5 | c.2479-1G>A | p.? | **6** | 226 | 4049 |  | rs775933506 |
|  | DNMT3A | NM_022552.5 | c.1667+1G>A | p.? | **2** | 63 | 3928 | COSM3720632 | rs776844136 |
| **060-RC** | PPM1D | NM_003620.4 | c.1547C>G | p.(Ser516*) | **2** | 161 | 9050 | COSM4423003 | rs772936724 |
|  | ZRSR2 | NM_005089.4 | c.524A>G | p.(Tyr175Cys) | **14** | 398 | 2830 |  |  |
| **065-MM** | DNMT3A | NM_022552.5 | c.2389A>G | p.(Asn797Asp) | **2** | 31 | 1397 | COSM5878752 |  |
|  | DNMT3A | NM_022552.5 | c.2114T>C | p.(Ile705Thr) | **8** | 245 | 2908 | COSM1583102 | rs777037011 |
| **066-NG** | DNMT3A | NM_022552.5 | c.2711C>T | p.(Pro904Leu) | **1** | 20 | 1910 | COSM87007 | rs149095705 |
| **067-PL** | DNMT3A | NM_022552.5 | c.1903C>T | p.(Arg635Trp) | **40** | 893 | 2223 | COSM87012 | rs144689354 |
|  | JAK2 | NM_004972.4 | c.1849G>T | p.(Val617Phe) | **1** | 37 | 2694 | COSM12600 | rs77375493 |
| **069-EU** | TET2 | NM_001127208.3 | c.5698G>T | p.(Val1900Phe) | **2** | 60 | 3723 | COSM4766132 |  |
| **070-GD** | TET2 | NM_001127208.3 | c.5609C>G | p.(Ser1870*) | **1** | 36 | 2883 | COSM6907542 |  |
| **071-AT** | TET2 | NM_001127208.3 | c.1795del | p.(Gln599Lysfs*2) | **1** | 57 | 3825 |  |  |
|  | TET2 | NM_001127208.3 | c.4097G>T | p.(Arg1366Leu) | **1** | 30 | 2622 |  |  |
| **073-GG** | DNMT3A | NM_022552.5 | c.2580G>A | p.(Trp860*) | **4** | 64 | 1778 | COSM4169946 | rs376830288 |
|  | TET2 | NM_001127208.3 | c.1496del | p.(Pro499Hisfs*34) | **5** | 197 | 4307 | COSM211641 |  |
|  | ASXL1 | NM_015338.6 | c.1210C>T | p.(Arg404*) | **5** | 126 | 2502 | COSM41598 | rs373145711 |
| **074-AL** | DNMT3A | NM_022552.5 | c.2644C>T | p.(Arg882Cys) | **3** | 63 | 1951 | COSM53042 | rs377577594 |
|  | DNMT3A | NM_022552.5 | c.1062_1063del | p.(Phe354Leufs*38) | **5** | 124 | 2531 |  |  |
|  | TET2 | NM_001127208.3 | c.4145A>G | p.(His1382Arg) | **4** | 73 | 1834 | COSM4605321 |  |
| **075-MH** | TET2 | NM_001127208.3 | c.2010_2011insT | p.(Ala671Cysfs*10) | **1** | 59 | 4796 |  |  |
|  | TET2 | NM_001127208.3 | c.4546C>T | p.(Arg1516*) | **1** | 28 | 2036 | COSM43420 | rs370735654 |
|  | TET2 | NM_001127208.3 | c.4615C>T | p.(Gln1539*) | **2** | 55 | 3176 | COSM144515 |  |
|  | SRSF2 | NM_003016.4 | c.283C>G | p.(Pro95Ala) | **13** | 208 | 1588 | COSM307352 | rs752405402 |
|  | SMC1A | NM_006306.4 | c.2330T>C | p.(Phe777Ser) | **1** | 25 | 2235 |  |  |
| **078-MM** | DNMT3A | NM_022552.5 | c.2255_2257del | p.(Phe752del) | **7** | 206 | 2843 |  | rs749132507 |
|  | TET2 | NM_001127208.3 | c.4067_4071dup | p.(Cys1358Glnfs*7) | **21** | 469 | 2212 |  |  |
| **079-PV** | TET2 | NM_001127208.3 | c.3851C>T | p.(Ser1284Phe) | **1** | 42 | 3300 | COSM120177 |  |
|  | U2AF1 | NM_006758.3 | c.470A>C | p.(Gln157Pro) | **1** | 58 | 5061 | COSM211534 | rs371246226 |
| **080-SH** | DNMT3A | NM_022552.5 | c.2333T>G | p.(Val778Gly) | **6** | 104 | 1602 |  | rs979932565 |
|  | DNMT3A | NM_022552.5 | c.2063G>A | p.(Arg688His) | **17** | 544 | 3189 | COSM4169953 | rs369713081 |
| **081-GC** | TET2 | NM_001127208.3 | c.2713del | p.(Asp905Ilefs*16) | 1 | 56 | 4111 |  |  |
|  | ASXL1 | NM_015338.6 | c.1249C>T | p.(Arg417*) | 5 | 130 | 2849 | COSM2270982 | rs375215583 |
| **082-MF** | TET2 | NM_001127208.3 | c.3594+1G>A | p.? | **3** | 98 | 2843 |  |  |
| **083-IB** | DNMT3A | NM_022552.5 | c.2200T>G | p.(Phe734Val) | **8** | 246 | 3005 | COSM7325450 | rs751950768 |
|  | SF3B1 | NM_012433.4 | c.2098A>G | p.(Lys700Glu) | **2** | 68 | 3744 | COSM84677 | rs559063155 |
| **084-GG** | ASXL1 | NM_015338.6 | c.1249C>T | p.(Arg417*) | **1** | 72 | 5498 | COSM2270982 | rs375215583 |
| **087-JM** | TET2 | NM_001127208.3 | c.1705dup | p.(Ala569Glyfs*14) | **5** | 274 | 5848 |  |  |
| **090-AD** | SF3B1 | NM_012433.4 | c.1997A>T | p.(Lys666Met) | **3** | 182 | 5714 | COSM110698 | rs374250186 |
| **091-MV** | DNMT3A | NM_022552.5 | c.2723A>G | p.(Tyr908Cys) | **2** | 69 | 3107 | COSM9351515 | rs780666472 |
|  | SF3B1 | NM_012433.4 | c.2098A>G | p.(Lys700Glu) | **1** | 36 | 3056 | COSM84677 | rs559063155 |
|  | SH2B3 | NM_005475.3 | c.1100G>A | p.(Gly367Asp) | **1** | 46 | 3868 | COSM6935448 |  |
| **094-MO** | DNMT3A | NM_022552.5 | c.1241dup | p.(Gln415Profs*6) | **8** | 157 | 1873 |  |  |
|  | TET2 | NM_001127208.3 | c.3405T>G | p.(Cys1135Trp) | **1** | 23 | 1891 |  |  |
| **097-LB** | DNMT3A | NM_022552.5 | c.918G>T | p.(Trp306Cys) | **9** | 438 | 4692 |  |  |
|  | TET2 | NM_001127208.3 | c.5698G>A | p.(Val1900Ile) | **3** | 169 | 5868 | COSM6996885 |  |
|  | TP53 | NM_001126112.3 | c.559+1G>A | p.? | **1** | 41 | 3667 | COSM6901 |  |
| **100-YP** | DNMT3A | NM_022552.5 | c.2711C>T | p.(Pro904Leu) | 4 | 199 | 4849 | COSM87007 | rs149095705 |
| **103-AL** | TET2 | NM_001127208.3 | c.5088T>A | p.(Tyr1696*) | **1** | 127 | 11139 |  |  |
| **104-PA** | TP53 | NM_001126112.3 | c.645T>G | p.(Ser215Arg) | **2** | 73 | 3757 | COSM44979 | rs1057520001 |
| **105-PN** | TET2 | NM_001127208.3 | c.5518_5542del | p.(Ala1840Glnfs*39) | **15** | 1152 | 7574 |  |  |
|  | PPM1D | NM_003620.4 | c.1349dup | p.(Leu450Phefs*6) | **1** | 47 | 4257 | COSM8946569 |  |
| **106-MM** | DNMT3A | NM_022552.5 | c.2408G>A | p.(Arg803Lys) | **1** | 40 | 2688 | COSM5944901 | rs764146514 |
| **107-ML** | DNMT3A | NM_022552.5 | c.2394del | p.(Gly800Valfs*2) | 2 | 25 | 1564 |  |  |
|  | TET2 | NM_001127208.3 | c.3081dup | p.(Met1028Hisfs*15) | 2 | 164 | 7014 |  |  |
| **108-MB** | U2AF1 | NM_006758.3 | c.101C>T | p.(Ser34Phe) | **12** | 600 | 4949 | COSM166866 | rs371769427 |
| **109-SC** | DNMT3A | NM_022552.5 | c.901C>T | p.(Arg301Trp) | **1** | 61 | 5276 | COSM2911819 |  |
|  | PPM1D | NM_003620.4 | c.1434C>A | p.(Cys478*) | **2** | 151 | 7061 | COSM98068 | rs146477590 |
| **111-BD** | TET2 | NM_001127208.3 | c.5633G>C | p.(Arg1878Pro) | **1** | 72 | 6746 |  |  |
| **112-JV** | STAG2 | NM_001042749.2 | c.3134G>A | p.(Arg1045Gln) | **3** | 155 | 6050 | COSM1227726 |  |
| **GA-007** | DNMT3A | NM_022552.5 | c.2645G>A | p.(Arg882His) | **1** | 21 | 1875 | COSM52944 | rs147001633 |
| **MB - 042** | DNMT3A | NM_022552.5 | c.1517A>T | p.(His506Leu) | **4** | 99 | 2641 |  |  |
| **JB - 038** | DNMT3A | NM_022552.5 | c.2651C>T | p.(Ala884Val) | **2** | 57 | 2356 | COSM231547 | rs559023562 |
|  | DNMT3A | NM_022552.5 | c.2335A>G | p.(Met779Val) | **1** | 12 | 1084 |  |  |
|  | DNMT3A | NM_022552.5 | c.1904G>A | p.(Arg635Gln) | **1** | 31 | 2751 | COSM1583088 | rs751562376 |
| **AM-060** | DNMT3A | NM_022552.5 | c.2578T>C | p.(Trp860Arg) | **26** | 698 | 2651 | COSM231568 | rs373014701 |
|  | TET2 | NM_001127208.3 | c.3804-2A>G | p.? | **9** | 210 | 2244 | COSM5879023 | rs576934285 |
|  | SAMD9 | NM_017654.4 | c.730_734del | p.(His244Glufs*10) | **2** | 107 | 5128 |  |  |
| **CC-028** | TET2 | NM_001127208.3 | c.3426del | p.(Asp1143Metfs*9) | **2** | 22 | 1001 | COSM1168034 |  |
| **CP-030** | DNMT3A | NM_022552.5 | c.1667+1G>A | p.? | **10** | 148 | 1557 | COSM3720632 | rs776844136 |
|  | ASXL1 | NM_015338.6 | c.1934dup | p.(Gly646Trpfs*12) | **4** | 106 | 3002 | COSM34210 |  |
| **GM-024** | DNMT3A | NM_022552.5 | c.2323-1G>A | p.? | **1** | 18 | 1241 |  |  |
| **RL-014** | DNMT3A | NM_022552.5 | c.2597+2T>G | p.? | **2** | 34 | 1605 | COSM53099 |  |
|  | TET2 | NM_001127208.3 | c.1157dup | p.(Phe387Valfs*3) | **1** | 24 | 1984 |  |  |
| **PE-017** | PPM1D | NM_003620.4 | c.1615G>T | p.(Glu539*) | **2** | 81 | 4452 |  |  |
| **JN-018** | TET2 | NM_001127208.3 | c.822del | p.(Asn275Ilefs*18) | **1** | 53 | 4374 | COSM43437 | rs777145283 |
|  | TET2 | NM_001127208.3 | c.1388del | p.(Pro463Leufs*23) | **9** | 252 | 2930 | COSM7314904 |  |
|  | TET2 | NM_001127208.3 | c.3594+2T>C | p.? | **6** | 73 | 1293 |  |  |
|  | TET2 | NM_001127208.3 | c.3965T>A | p.(Leu1322Gln) | **30** | 501 | 1681 | COSM211728 | rs551797762 |
|  | TET2 | NM_001127208.3 | c.4076G>A | p.(Arg1359His) | **2** | 40 | 2292 | COSM42055 | rs775677220 |
|  | TP53 | NM_001126112.3 | c.658del | p.(Tyr220Metfs*27) | **3** | 42 | 1627 | COSM6008894 |  |
|  | PPM1D | NM_003620.4 | c.1654C>T | p.(Arg552*) | **0.9** | 21 | 2253 | COSM982226 | rs779070661 |
|  | SRSF2 | NM_003016.4 | c.284C>A | p.(Pro95His) | **21** | 296 | 1414 | COSM211029,COSM211504 | rs751713049 |
| **JF-022** | DNMT3A | NM_022552.5 | c.2115del | p.(Ile705Metfs*74) | **8** | 179 | 2148 |  |  |
|  | ASXL2 | NM_018263.6 | c.1966_1979del | p.(Thr656Glnfs*46) | **1** | 21 | 1731 |  |  |
|  | JAK2 | NM_004972.4 | c.1711G>A | p.(Gly571Ser) | **40** | 458 | 1146 | COSM29107 | rs139504737 |
| **TP-013** | TET2 | NM_001127208.3 | c.3313A>G | p.(Ile1105Val) | **3** | 99 | 3785 |  |  |
|  | IKZF1 | NM_006060.6 | c.476A>G | p.(Asn159Ser) | **1** | 55 | 4549 | COSM5487593 | rs374333820 |
|  | KRAS | NM_033360.4 | c.38G>A | p.(Gly13Asp) | **1** | 40 | 3294 | COSM532 | rs112445441 |
| **MD-012** | ZRSR2 | NM_005089.4 | c.1030A>G | p.(Arg344Gly) | **1** | 42 | 3296 | COSM6023688 |  |
| **MC-031** | TET2 | NM_001127208.3 | c.2290dup | p.(Gln764Profs*5) | **4** | 133 | 3674 | COSM43504 |  |
| **BT-016** | DNMT3A | NM_175629.2 | c.1702G>C | p.(Gly568Arg) | **1** | 21 | 1643 |  |  |
| **FD-033** | DNMT3A | NM_175629.2 | c.2478+1G>C | p.? | **1** | 12 | 970 |  |  |
| **FF-032** | DNMT3A | NM_175629.2 | c.1430-2A>G | p.? | **1** | 30 | 2517 |  |  |
|  | TP53 | NM_000546.6 | c.646G>A | p.(Val216Met) | **1** | 26 | 2318 | COSM10667 | rs730882025 |
|  | TP53 | NM_000546.6 | c.358A>G | p.(Lys120Glu) | **1** | 20 | 1511 | COSM44827 | rs121912658 |
| **JL-034** | PPM1D | NM_003620.4 | c.1636dup | p.(Leu546Profs*6) | **1** | 43 | 3121 | COSM7346322 |  |
| **PR-036** | DNMT3A | NM_175629.2 | c.1015-1G>A | p.? | **1** | 12 | 828 |  |  |
| **SH-037** | DNMT3A | NM_175629.2 | c.2119G>A | p.(Gly707Ser) | **1** | 14 | 1235 | COSM7409654 |  |
|  | DNMT3A | NM_175629.2 | c.1979A>G | p.(Tyr660Cys) | **1** | 35 | 3138 | COSM256036 | rs767552800 |
| **JC-039** | PPM1D | NM_003620.4 | c.1422del | p.(Glu475Lysfs*8) | **10** | 238 | 2452 |  |  |
|  | ASXL1 | NM_015338.6 | c.2074C>T | p.(Gln692*) | **9** | 203 | 2293 | COSM1155830 |  |
|  | U2AF1 | NM_001025203.1 | c.22A>G | p.(Ile8Val) | **1** | 18 | 1747 |  |  |
| **PG-040** | DNMT3A | NM_175629.2 | c.2645G>A | p.(Arg882His) | **2** | 26 | 1575 | COSM52944 | rs147001633 |
| **CM-045** | DNMT3A | NM_175629.2 | c.811dup | p.(Asp271Glyfs*10) | **4** | 40 | 891 |  |  |
|  | TET2 | NM_001127208.3 | c.822del | p.(Asn275Ilefs*18) | **5** | 141 | 2850 | COSM43437 | rs777145283 |
| **LB-046** | DNMT3A | NM_175629.2 | c.2372C>A | p.(Ala791Asp) | **1** | 9 | 646 |  |  |
|  | DNMT3A | NM_175629.2 | c.2204A>C | p.(Tyr735Ser) | **1** | 13 | 1033 | COSM4908593 | rs147828672 |
|  | PPM1D | NM_003620.4 | c.1508C>G | p.(Ser503*) | **2** | 55 | 3541 |  |  |
|  | ASXL1 | NM_015338.6 | c.1934dup | p.(Gly646Trpfs*12) | **2** | 41 | 2175 | COSM34210 |  |
|  | PHF6 | NM_001015877.2 | c.1001A>G | p.(Glu334Gly) | **1** | 17 | 1596 |  |  |
| **MS-006** | DNMT3A | NM_022552.5 | c.2264T>C | p.(Phe755Ser) | **1** | 74 | 5525 | COSM2911748 | rs536841393 |
|  | TET2 | NM_001127208.3 | c.5038C>T | p.(Gln1680*) | **4** | 350 | 8201 | COSM132898 | rs997900764 |
|  | ASXL1 | NM_015338.6 | c.1636C>T | p.(Gln546*) | **12** | 454 | 3807 | COSM5989986 |  |

**Supplementary Table S4. Mutational profile of CHIP(+) MI(-) subjects**

| **Patient** | **Gene** | **Transcript** | **c.** | **p.** | **Freq** | **Var.Cov.** | **Depth** | **COSMIC** | **dbSNP** |
| --- | --- | --- | --- | --- | --- | --- | --- | --- | --- |
| **20606** | ASXL2 | NM_018263.6 | c.1379C>T | p.(Ser460Phe) | 2 | 78 | 4680 |  | rs750947456 |
| **20634** | DNMT3A | NM_022552.5 | c.2598-1G>C | p.? | 1 | 18 | 1330 |  | rs766506181 |
|  | DNMT3A | NM_022552.5 | c.2339T>C | p.(Ile780Thr) | 7 | 83 | 1206 | COSM1583121 | rs370751539 |
|  | DNMT3A | NM_022552.5 | c.2311C>T | p.(Arg771*) | 8 | 141 | 1791 | COSM231563 | rs779626155 |
|  | DNMT3A | NM_022552.5 | c.2120G>A | p.(Gly707Asp) | 2 | 53 | 2153 | COSM5574184 | rs927337009 |
| **20646** | TET2 | NM_001127208.3 | c.4870C>T | p.(Gln1624*) | 1 | 61 | 4475 | COSM42020 |  |
| **20767** | TET2 | NM_001127208.3 | c.1632del | p.(Asp545Thrfs*16) | 1 | 63 | 5694 |  |  |
| **20774** | DNMT3A | NM_022552.5 | c.1443C>G | p.(Tyr481*) | 2 | 63 | 3837 |  |  |
| **20785** | ZRSR2 | NM_005089.4 | c.524A>G | p.(Tyr175Cys) | 3 | 40 | 1163 |  |  |
| **20809** | DNMT3A | NM_022552.5 | c.980G>A | p.(Trp327*) | 1 | 71 | 5553 | COSM6907493 | rs750966422 |
| **20841** | TET2 | NM_001127208.3 | c.5603_5604del | p.(His1868Argfs*6) | 6 | 255 | 4485 |  |  |
| **20845** | DNMT3A | NM_022552.5 | c.1180dup | p.(Asp394Glyfs*15) | 10 | 158 | 1628 |  |  |
|  | TP53 | NM_001126112.3 | c.239C>G | p.(Pro80Arg) | 6 | 215 | 3622 |  |  |
| **21118** | TET2 | NM_001127208.3 | c.1771C>T | p.(Gln591*) | 4 | 130 | 3329 | COSM42052 |  |
|  | IDH2 | NM_002168.4 | c.419G>A | p.(Arg140Gln) | 8 | 180 | 2302 | COSM41590 | rs121913502 |
|  | SRSF2 | NM_003016.4 | c.284_304del | p.(Pro95_Ser101del) | 17 | 258 | 1481 |  |  |
| **21138** | SF3B1 | NM_012433.4 | c.1997A>G | p.(Lys666Arg) | 7 | 254 | 3755 | COSM131553 | rs374250186 |
|  | ASXL1 | NM_015338.6 | c.2389G>T | p.(Glu797*) | 2 | 117 | 7789 | COSM303906 |  |
|  | ASXL1 | NM_015338.6 | c.2461del | p.(Asp821Ilefs*3) | 1 | 65 | 5129 |  |  |
| **21184** | SF3B1 | NM_012433.4 | c.2098A>G | p.(Lys700Glu) | 1 | 48 | 3836 | COSM84677 | rs559063155 |
| **21238** | TET2 | NM_001127208.3 | c.3374del | p.(Lys1125Argfs*12) | 1 | 23 | 1836 |  |  |
| **21244** | DNMT3A | NM_022552.5 | c.2339T>C | p.(Ile780Thr) | 1 | 20 | 1570 | COSM1583121 | rs370751539 |
|  | DNMT3A | NM_022552.5 | c.1598A>G | p.(Tyr533Cys) | 2 | 81 | 5367 | COSM231555 |  |
| **21262** | TET2 | NM_001127208.3 | c.4127A>G | p.(Asp1376Gly) | 3 | 113 | 3821 |  |  |
| **21378** | DNMT3A | NM_022552.5 | c.1429+1G>T | p.? | 2 | 78 | 4203 |  |  |
| **20850** | DNMT3A | NM_022552.5 | c.1516_1517insG | p.(His506Argfs*40) | 2 | 55 | 2275 | COSM6907582 |  |
| **20868** | STAG2 | NM_001042749.2 | c.2864T>G | p.(Phe955Cys) | 1 | 19 | 1808 |  |  |
| **20912** | TET2 | NM_001127208.3 | c.2132del | p.(Glu711Glyfs*40) | 25 | 1299 | 5241 |  |  |
| **20945** | DNMT3A | NM_022552.5 | c.976C>T | p.(Arg326Cys) | 26 | 897 | 3505 | COSM4169721 | rs747448117 |
|  | TET2 | NM_001127208.3 | c.840dup | p.(Asn281*) | 24 | 1321 | 5394 | COSM100073 |  |
|  | TET2 | NM_001127208.3 | c.4438del | p.(Ser1480Profs*91) | 27 | 1119 | 4178 |  |  |
|  | SH2B3 | NM_005475.3 | c.1241T>G | p.(Leu414Arg) | 4 | 220 | 4978 |  |  |
|  | U2AF1 | NM_006758.3 | c.101C>T | p.(Ser34Phe) | 8 | 224 | 2877 | COSM166866 | rs371769427 |
| **20990** | TET2 | NM_001127208.3 | c.3621G>C | p.(Glu1207Asp) | 8 | 196 | 2540 |  |  |
|  | ASXL1 | NM_015338.6 | c.1934dup | p.(Gly646Trpfs*12) | 21 | 779 | 3690 | COSM34210 |  |
| **21031** | TET2 | NM_001127208.3 | c.3578G>A | p.(Cys1193Tyr) | 1 | 20 | 1632 | COSM1737882 |  |
| **21047** | JAK2 | NM_004972.4 | c.1849G>T | p.(Val617Phe) | 2 | 40 | 1934 | COSM12600 | rs77375493 |
| **21057** | RUNX1 | NM_001754.5 | c.171_198del | p.(Leu58Trpfs*5) | 2 | 143 | 6561 |  |  |
| **21092** | GNAS | NM_000516.7 | c.602G>A | p.(Arg201His) | 1 | 18 | 1522 | COSM27895 | rs121913495 |
| **21167** | DNMT3A | NM_022552.5 | c.1551C>G | p.(Cys517Trp) | 1 | 51 | 4392 |  |  |
| **21387** | CUX1 | NM_181552.4 | c.1234_1235insTTGT | p.(Arg412Ilefs*9) | 3 | 73 | 2756 |  |  |
| **21401** | DNMT3A | NM_022552.5 | c.2508del | p.(Arg836Serfs*5) | 1 | 22 | 2063 |  |  |
| **21481** | U2AF1 | NM_006758.3 | c.467G>A | p.(Arg156His) | 9 | 543 | 6279 | COSM1235014 | rs769567889 |
| **21484** | DNMT3A | NM_022552.5 | c.1084C>T | p.(Gln362*) | 2 | 84 | 5291 | COSM6907477 |  |
|  | TET2 | NM_001127208.3 | c.4317dup | p.(Arg1440Thrfs*38) | 1 | 56 | 4492 | COSM4385603 |  |
| **21507** | DNMT3A | NM_022552.5 | c.2309C>T | p.(Ser770Leu) | 2 | 87 | 3651 | COSM231549 | rs758845779 |
| **21508** | DNMT3A | NM_022552.5 | c.2086C>T | p.(Gln696*) | 13 | 583 | 4479 |  | rs750325978 |
|  | DNMT3A | NM_022552.5 | c.884T>C | p.(Leu295Pro) | 2 | 72 | 3681 | COSM6908459 |  |
|  | TET2 | NM_001127208.3 | c.3581del | p.(Pro1194Leufs*32) | 8 | 188 | 2422 |  |  |
| **21536** | TET2 | NM_001127208.3 | c.4534G>A | p.(Ala1512Thr) | 14 | 467 | 3455 |  |  |
|  | CBL | NM_005188.4 | c.1139T>C | p.(Leu380Pro) | 2 | 40 | 2525 | COSM34055 |  |
| **21547** | DNMT3A | NM_022552.5 | c.2728G>C | p.(Ala910Pro) | 4 | 149 | 3821 | COSM4383517 |  |
| **21642** | TET2 | NM_001127208.3 | c.5180_5181del | p.(His1727Argfs*12) | 1 | 57 | 5396 |  |  |
| **21663** | DNMT3A | NM_022552.5 | c.2339T>C | p.(Ile780Thr) | 2 | 44 | 2451 | COSM1583121 | rs370751539 |
|  | DNMT3A | NM_022552.5 | c.1488del | p.(Cys497Valfs*154) | 6 | 228 | 3981 |  |  |
| **21673** | DNMT3A | NM_022552.5 | c.1430-2A>G | p.? | 1 | 70 | 6239 |  |  |
| **21690** | DNMT3A | NM_022552.5 | c.2679G>C | p.(Trp893Cys) | 14 | 584 | 4057 | COSM1583151 |  |
| **21714** | DNMT3A | NM_022552.5 | c.2339T>C | p.(Ile780Thr) | 2 | 58 | 2758 | COSM1583121 | rs370751539 |
|  | KIT | NM_000222.3 | c.2752A>G | p.(Lys918Glu) | 4 | 197 | 5307 |  |  |
|  | BCORL1 | NM_021946.5 | c.3977T>G | p.(Leu1326Arg) | 2 | 18 | 941 |  |  |
| **21730** | CSF3R | NM_156039.3 | c.815C>T | p.(Pro272Leu) | 13 | 343 | 2708 |  | rs367774391 |
|  | DNMT3A | NM_022552.5 | c.2479-1G>C | p.? | 12 | 369 | 3186 |  |  |
|  | TET2 | NM_001127208.3 | c.3409+1G>A | p.? | 3 | 64 | 2009 |  | rs772634266 |
|  | TET2 | NM_001127208.3 | c.5666C>G | p.(Pro1889Arg) | 12 | 607 | 5092 | COSM211750 |  |
| **21740** | ASXL1 | NM_015338.6 | c.2060_2061del | p.(Cys687Tyrfs*30) | 9 | 548 | 6307 | COSM146261 |  |
| **21755** | DNMT3A | NM_022552.5 | c.2520_2525delinsGAAGCG | p.(Ile840_Gln842delinsMetLysArg) | 20 | 816 | 4005 |  |  |
| **21757** | DNMT3A | NM_022552.5 | c.2185C>T | p.(Arg729Trp) | 4 | 105 | 2607 | COSM249142 | rs200018028 |
|  | DNMT3A | NM_022552.5 | c.1374del | p.(Lys459Argfs*192) | 1 | 53 | 5067 |  | rs886041641 |
| **21793** | DNMT3A | NM_022552.5 | c.1279+1G>A | p.? | 2 | 21 | 1211 | COSM6938921 | rs374440649 |
| **21830** | IDH1 | NM_005896.4 | c.311G>T | p.(Gly104Val) | 2 | 22 | 1227 |  |  |
|  | IDH1 | NM_005896.4 | c.155C>A | p.(Thr52Asn) | 5 | 33 | 702 |  |  |
|  | SMC3 | NM_005445.4 | c.3149G>T | p.(Gly1050Val) | 9 | 72 | 769 |  |  |
|  | SMC3 | NM_005445.4 | c.3644C>A | p.(Thr1215Lys) | 2 | 16 | 662 |  |  |
|  | KRAS | NM_033360.4 | c.35G>T | p.(Gly12Val) | 3 | 26 | 747 | COSM520 | rs121913529 |
|  | ASXL1 | NM_015338.6 | c.2122C>T | p.(Gln708*) | 14 | 822 | 5701 | COSM1234988 | rs755789372 |
| **21836** | DNMT3A | NM_022552.5 | c.1906G>A | p.(Val636Met) | 3 | 61 | 1902 | COSM133124 | rs376550450 |
|  | DNMT3A | NM_022552.5 | c.1652dup | p.(Asn551Lysfs*27) | 2 | 32 | 1610 |  |  |
|  | SF3B1 | NM_012433.4 | c.2098A>G | p.(Lys700Glu) | 3 | 53 | 1608 | COSM84677 | rs559063155 |
|  | PPM1D | NM_003620.4 | c.1349del | p.(Leu450*) | 1 | 24 | 2024 | COSM2793542 |  |
| **21858** | DNMT3A | NM_022552.5 | c.2246G>A | p.(Arg749His) | 3 | 97 | 2877 | COSM6647818 | rs34843713 |
|  | DNMT3A | NM_022552.5 | c.2173+1G>A | p.? | 2 | 57 | 2283 | COSM6907619 | rs763716866 |
| **21888** | DNMT3A | NM_022552.5 | c.2204A>G | p.(Tyr735Cys) | 2 | 59 | 2896 | COSM133126 | rs147828672 |
| **21924** | DNMT3A | NM_022552.5 | c.2645G>A | p.(Arg882His) | 8 | 177 | 2266 | COSM52944 | rs147001633 |
| **22283** | DNMT3A | NM_022552.5 | c.2204A>G | p.(Tyr735Cys) | 3 | 81 | 2610 | COSM133126 | rs147828672 |
| **21598** | TP53 | NM_001126112.3 | c.734G>A | p.(Gly245Asp) | 1 | 10 | 683 | COSM43606 | rs121912656 |
| **21605** | DNMT3A | NM_022552.5 | c.2173+1G>A | p.? | 4 | 44 | 1155 | COSM6907619 | rs763716866 |
|  | TET2 | NM_001127208.3 | c.3699G>A | p.(Trp1233*) | 13 | 240 | 1794 | COSM211667 |  |
|  | TET2 | NM_001127208.3 | c.3954+1G>A | p.? | 1 | 13 | 1104 | COSM87141 | rs1011965788 |
|  | TET2 | NM_001127208.3 | c.4139A>G | p.(His1380Arg) | 2 | 21 | 1121 | COSM2270975 |  |
|  | U2AF1 | NM_006758.3 | c.470A>G | p.(Gln157Arg) | 11 | 291 | 2744 | COSM211532 | rs371246226 |
| **22244** | DNMT3A | NM_022552.5 | c.1906G>A | p.(Val636Met) | 7 | 82 | 1135 | COSM133124 | rs376550450 |
|  | ZRSR2 | NM_005089.4 | c.1307_1312delinsACC | p.(Gly436_Gly438delinsAspArg) | 8 | 105 | 1268 |  |  |
| **22256** | TET2 | NM_001127208.3 | c.1842dup | p.(Leu615Alafs*23) | 13 | 620 | 4826 | COSM43478 |  |
| **22407** | DNMT3A | NM_022552.5 | c.2477A>G | p.(Lys826Arg) | 1 | 21 | 1755 | COSM1583124 | rs770079872 |
| **22501** | ASXL1 | NM_015338.6 | c.1934dup | p.(Gly646Trpfs*12) | 9 | 255 | 2843 | COSM34210 |  |
| **21611** | DNMT3A | NM_022552.5 | c.2026C>T | p.(Arg676Trp) | 5 | 178 | 3932 | COSM328646 | rs375399431 |
| **21899** | DNMT3A | NM_022552.5 | c.1490G>A | p.(Cys497Tyr) | 2 | 83 | 3850 | COSM1318925 | rs779323387 |
| **21953** | DNMT3A | NM_022552.5 | c.1795G>T | p.(Glu599*) | 2 | 111 | 5601 |  |  |
| **21998** | SMC3 | NM_005445.4 | c.3149G>T | p.(Gly1050Val) | 1 | 35 | 3335 |  |  |
|  | SH2B3 | NM_005475.3 | c.914G>A | p.(Cys305Tyr) | 24 | 1590 | 6646 |  | rs756955512 |
| **22039** | TET2 | NM_001127208.3 | c.2263G>T | p.(Glu755*) | 2 | 106 | 6789 | COSM4766087 |  |
| **22081** | DNMT3A | NM_022552.5 | c.2645G>A | p.(Arg882His) | 41 | 2027 | 4885 | COSM52944 | rs147001633 |
|  | TET2 | NM_001127208.3 | c.3851C>T | p.(Ser1284Phe) | 1 | 64 | 5364 | COSM120177 |  |
|  | TET2 | NM_001127208.3 | c.5673G>T | p.(Arg1891Ser) | 2 | 159 | 7312 |  |  |
| **22097** | EZH2 | NM_004456.5 | c.1803del | p.(Ser601Argfs*74) | 1 | 16 | 1247 |  |  |
|  | FLT3 | NM_004119.3 | c.882+2del | p.? | 1 | 17 | 1183 |  |  |
|  | BCOR | NM_001123385.2 | c.4442A>T | p.(Tyr1481Phe) | 1 | 26 | 1740 |  |  |
|  | STAG2 | NM_001042749.2 | c.1297T>A | p.(Tyr433Asn) | 1 | 13 | 1050 |  |  |
| **24389** | DNMT3A | NM_022552.5 | c.1554+1G>A | p.? | 1 | 34 | 2853 | COSM4775128 | rs766110518 |
|  | TET2 | NM_001127208.3 | c.3893G>A | p.(Cys1298Tyr) | 6 | 325 | 5612 | COSM87138 |  |
| **24410** | PHF6 | NM_001015877.2 | c.277G>C | p.(Gly93Arg) | 4 | 108 | 2716 |  |  |
| **24422** | RAD21 | NM_006265.3 | c.807T>G | p.(Asn269Lys) | 2 | 59 | 2942 |  |  |
| **22510** | DNMT3A | NM_022552.5 | c.2206C>T | p.(Arg736Cys) | 2 | 75 | 4981 | COSM231560 | rs761934754 |
| **22511** | DNMT3A | NM_022552.5 | c.890G>C | p.(Trp297Ser) | 1 | 65 | 5202 |  |  |
|  | IDH1 | NM_005896.4 | c.155C>A | p.(Thr52Asn) | 2 | 39 | 2039 |  |  |
|  | KIT | NM_000222.3 | c.269C>A | p.(Thr90Asn) | 2 | 63 | 3429 |  |  |
| **22591** | SH2B3 | NM_005475.3 | c.1117A>T | p.(Lys373*) | 2 | 95 | 5693 |  |  |
| **22603** | DNMT3A | NM_022552.5 | c.1123-2A>C | p.? | 2 | 44 | 2029 |  | rs752605931 |
| **22633** | ASXL1 | NM_015338.6 | c.1438G>T | p.(Glu480*) | 13 | 986 | 7538 | COSM3719083 | rs545224250 |
| **22643** | TP53 | NM_001126112.3 | c.659A>G | p.(Tyr220Cys) | 4 | 130 | 3556 | COSM10758 | rs121912666 |
| **22704** | TET2 | NM_001127208.3 | c.4664_4665del | p.(Glu1555Valfs*22) | 3 | 127 | 5057 | COSM4383953 |  |
|  | ASXL1 | NM_015338.6 | c.1534C>T | p.(Gln512*) | 3 | 186 | 6210 | COSM443634 | rs757832294 |
| **22858** | TET2 | NM_001127208.3 | c.1299_1300insG | p.(His434Alafs*9) | 1 | 81 | 5582 |  |  |
|  | TET2 | NM_001127208.3 | c.4552del | p.(Ser1518Glnfs*53) | 3 | 80 | 2926 |  |  |
| **22862** | TET2 | NM_001127208.3 | c.987dup | p.(Glu330*) | 3 | 151 | 5957 | COSM110781 |  |
| **22874** | DNMT3A | NM_022552.5 | c.2058C>A | p.(Asp686Glu) | 17 | 663 | 3918 |  |  |
| **22892** | DNMT3A | NM_022552.5 | c.1453C>T | p.(Gln485*) | 22 | 749 | 3405 | COSM6907791 |  |
|  | DNMT3A | NM_022552.5 | c.1430-1G>C | p.? | 4 | 127 | 3367 |  |  |
| **22901** | DNMT3A | NM_022552.5 | c.2635A>G | p.(Asn879Asp) | 5 | 226 | 4872 | COSM1583135 |  |
|  | BCOR | NM_001123385.2 | c.3995T>C | p.(Val1332Ala) | 2 | 106 | 6098 |  |  |
| **22906** | DNMT3A | NM_022552.5 | c.890G>A | p.(Trp297*) | 6 | 266 | 4415 | COSM133740 | rs944608317 |
| **2300** | DNMT3A | NM_022552.5 | c.2363dup | p.(His789Thrfs*9) | 5 | 171 | 3195 |  |  |
| **23065** | EZH2 | NM_004456.5 | c.1876G>A | p.(Val626Met) | 4 | 132 | 3011 | COSM1000723 | rs587783625 |
|  | CBL | NM_005188.4 | c.1259G>A | p.(Arg420Gln) | 1 | 37 | 2775 | COSM34077 | rs267606708 |
|  | ASXL1 | NM_015338.6 | c.2324del | p.(Leu775*) | 34 | 1373 | 3984 | COSM53206 |  |
|  | U2AF1 | NM_006758.3 | c.470A>C | p.(Gln157Pro) | 2 | 69 | 3494 | COSM211534 | rs371246226 |
| **23075** | TET2 | NM_001127208.3 | c.1279_1282del | p.(Asn427Valfs*4) | 3 | 105 | 3765 | COSM6987213 |  |
| **23174** | DNMT3A | NM_022552.5 | c.2389A>G | p.(Asn797Asp) | 1 | 47 | 3623 | COSM5878752 |  |
| **24450** | DNMT3A | NM_022552.5 | c.2254T>C | p.(Phe752Leu) | 3 | 133 | 4950 | COSM133131 |  |
|  | TET2 | NM_001127208.3 | c.391dup | p.(Arg131Lysfs*5) | 1 | 104 | 8751 |  |  |
| **22753** | TET2 | NM_001127208.3 | c.1970C>G | p.(Ser657*) | 2 | 94 | 5205 | COSM4996387 |  |
| **22838** | DNMT3A | NM_022552.5 | c.2726T>G | p.(Phe909Cys) | 2 | 48 | 2453 | COSM166527 |  |
|  | DNMT3A | NM_022552.5 | c.2185C>T | p.(Arg729Trp) | 2 | 41 | 2576 | COSM249142 | rs200018028 |
|  | DNMT3A | NM_022552.5 | c.1994T>G | p.(Val665Gly) | 3 | 94 | 2896 | COSM231559 | rs762503226 |
| **22844** | DNMT3A | NM_022552.5 | c.2009_2011del | p.(Ile670del) | 2 | 75 | 3947 |  |  |
| **22847** | GNB1 | NM_002074.5 | c.169A>G | p.(Lys57Glu) | 3 | 49 | 1707 | COSM211450 | rs141326438 |
|  | DNMT3A | NM_022552.5 | c.1293del | p.(Tyr432Thrfs*219) | 1 | 13 | 1193 |  |  |
| **23313** | ASXL1 | NM_015338.6 | c.1282C>T | p.(Gln428*) | 11 | 308 | 2701 | COSM5945400 | rs886041975 |
|  | RUNX1 | NM_001754.5 | c.280A>T | p.(Ser94Cys) | 1 | 20 | 1577 |  |  |
| **23364** | DNMT3A | NM_022552.5 | c.1916T>A | p.(Leu639His) | 5 | 124 | 2326 |  |  |
|  | TET2 | NM_001127208.3 | c.1648C>T | p.(Arg550*) | 8 | 322 | 3821 | COSM41644 | rs572712965 |
|  | TET2 | NM_001127208.3 | c.3869C>A | p.(Ser1290*) | 9 | 283 | 3125 |  |  |
| **23368** | KRAS | NM_033360.4 | c.38G>A | p.(Gly13Asp) | 1 | 33 | 2757 | COSM532 | rs112445441 |
|  | ASXL1 | NM_015338.6 | c.1900_1922del | p.(Glu635Argfs*15) | 1 | 30 | 2239 | COSM36165 |  |
| **23461** | GNAS | NM_000516.7 | c.602G>A | p.(Arg201His) | 2 | 33 | 1385 | COSM27895 | rs121913495 |
| **23487** | SRSF2 | NM_003016.4 | c.284_307del | p.(Pro95_Arg102del) | 3 | 52 | 1716 |  | rs766200080 |
| **23274** | DNMT3A | NM_022552.5 | c.2402T>G | p.(Met801Arg) | 2 | 37 | 2414 |  | rs901395842 |
| **23509** | CSF3R | NM_156039.3 | c.1996G>A | p.(Ala666Thr) | 3 | 140 | 4669 |  |  |
| **23583** | EZH2 | NM_004456.5 | c.484+1G>A | p.? | 4 | 63 | 1418 |  |  |
|  | IDH2 | NM_002168.4 | c.418C>T | p.(Arg140Trp) | 22 | 515 | 2357 | COSM41877 | rs267606870 |
|  | SRSF2 | NM_003016.4 | c.284C>A | p.(Pro95His) | 20 | 363 | 1841 | COSM211029,COSM211504 | rs751713049 |
|  | ASXL1 | NM_015338.6 | c.1720-2A>G | p.? | 25 | 368 | 1490 |  | rs376029425 |
| **23591** | IDH1 | NM_005896.4 | c.155C>A | p.(Thr52Asn) | 1 | 35 | 2661 |  |  |
| **23604** | TET2 | NM_001127208.3 | c.358G>T | p.(Gly120*) | 7 | 517 | 7418 |  |  |
|  | SMC3 | NM_005445.4 | c.2597T>C | p.(Leu866Pro) | 3 | 99 | 3099 |  |  |
| **23700** | TET2 | NM_001127208.3 | c.3646C>T | p.(Arg1216*) | 1 | 101 | 7026 | COSM42029 | rs1009194427 |
|  | SRSF2 | NM_003016.4 | c.283C>A | p.(Pro95Thr) | 3 | 79 | 3018 | COSM307353 | rs752405402 |
| **23798** | SF3B1 | NM_012433.4 | c.1986C>A | p.(His662Gln) | 15 | 900 | 6173 | COSM130416 | rs778467242 |
|  | JAK2 | NM_004972.4 | c.1849G>T | p.(Val617Phe) | 4 | 171 | 4294 | COSM12600 | rs77375493 |
| **23806** | TET2 | NM_001127208.3 | c.3417del | p.(Ile1140Leufs*12) | 5 | 181 | 3296 |  |  |
|  | ASXL1 | NM_015338.6 | c.2519del | p.(Pro840Leufs*2) | 6 | 377 | 6351 |  |  |
| **23942** | DNMT3A | NM_022552.5 | c.977G>A | p.(Arg326His) | 1 | 71 | 5150 | COSM477213 | rs758881009 |
| **23986** | DNMT3A | NM_022552.5 | c.1752C>A | p.(Tyr584*) | 5 | 259 | 4740 | COSM5574186 |  |
|  | DNMT3A | NM_022552.5 | c.1628G>A | p.(Gly543Asp) | 11 | 427 | 4015 | COSM7409708 | rs767226511 |
| **24150** | PPM1D | NM_003620.4 | c.1372C>T | p.(Arg458*) | 3 | 164 | 4961 | COSM2793544 | rs773389405 |
| **24314** | DNMT3A | NM_022552.5 | c.2210T>A | p.(Leu737His) | 1 | 57 | 3889 | COSM133739 |  |
|  | DNMT3A | NM_022552.5 | c.1669T>G | p.(Cys557Gly) | 5 | 209 | 3938 |  | rs760791871 |
|  | TET2 | NM_001127208.3 | c.4567C>T | p.(Gln1523*) | 1 | 37 | 3611 | COSM96925 |  |
| **24315** | DNMT3A | NM_022552.5 | c.1136G>A | p.(Arg379His) | 1 | 30 | 2287 | COSM5772441 |  |
| **24319** | DNMT3A | NM_022552.5 | c.2262_2266del | p.(Phe755Glufs*8) | 2 | 98 | 4143 |  |  |
| **24326** | TET2 | NM_001127208.3 | c.3524T>G | p.(Ile1175Ser) | 4 | 96 | 2514 |  |  |
|  | TET2 | NM_001127208.3 | c.4022C>A | p.(Ala1341Glu) | 46 | 1194 | 2569 | COSM144544 |  |
|  | TET2 | NM_001127208.3 | c.5594_5603del | p.(Ala1865Valfs*19) | 4 | 180 | 4777 |  |  |
|  | CBL | NM_005188.4 | c.914T>C | p.(Ile305Thr) | 3 | 142 | 4072 |  |  |
|  | SRSF2 | NM_003016.4 | c.284C>G | p.(Pro95Arg) | 25 | 617 | 2442 | COSM211661 | rs751713049 |
|  | BCOR | NM_001123385.2 | c.652T>A | p.(Tyr218Asn) | 21 | 1350 | 6512 |  |  |
| **23059** | ASXL1 | NM_015338.6 | c.2206del | p.(Thr736Glnfs*8) | 3 | 153 | 5297 |  |  |
| **24496** | TET2 | NM_001127208.3 | c.4138C>T | p.(His1380Tyr) | 4 | 149 | 3700 | COSM87161 | rs577810844 |
|  | SAMD9 | NM_017654.4 | c.4581G>T | p.(Leu1527Phe) | 12 | 680 | 5746 |  |  |
| **24524** | DNMT3A | NM_022552.5 | c.1063del | p.(His355Thrfs*52) | 34 | 820 | 2385 |  |  |
| **24551** | DNMT3A | NM_022552.5 | c.1903C>T | p.(Arg635Trp) | 1 | 26 | 2499 | COSM87012 | rs144689354 |
| **24566** | TET2 | NM_001127208.3 | c.2402_2424del | p.(Asn801Serfs*7) | 3 | 136 | 4269 |  |  |
| **24587** | TET2 | NM_001127208.3 | c.685dup | p.(Thr229Asnfs*25) | 1 | 93 | 7925 | COSM110749 |  |
|  | KDM6A | NM_021140.4 | c.2437_2440del | p.(Asn813Leufs*53) | 5 | 164 | 3396 |  |  |
| **24617** | DNMT3A | NM_022552.5 | c.1585G>T | p.(Asp529Tyr) | 13 | 501 | 3878 |  |  |
|  | TET2 | NM_001127208.3 | c.3766G>A | p.(Gly1256Ser) | 46 | 1604 | 3522 |  |  |
| **24621** | TET2 | NM_001127208.3 | c.4305G>T | p.(Gln1435His) | 1 | 69 | 5621 | COSM1426218 |  |
| **24630** | DNMT3A | NM_022552.5 | c.2580G>A | p.(Trp860*) | 2 | 70 | 3326 | COSM4169946 | rs376830288 |
| **24689** | TET2 | NM_001127208.3 | c.5606G>A | p.(Gly1869Glu) | 6 | 227 | 3778 | COSM6963127 |  |
|  | ASXL1 | NM_015338.6 | c.1926dup | p.(Gly643Argfs*15) | 3 | 103 | 3928 | COSM307769 |  |
| **24722** | DNMT3A | NM_022552.5 | c.2732G>A | p.(Cys911Tyr) | 1 | 24 | 2079 | COSM9277296 | rs906113912 |
|  | DNMT3A | NM_022552.5 | c.1637T>G | p.(Val546Gly) | 2 | 89 | 3719 |  |  |
| **24723** | DNMT3A | NM_022552.5 | c.2645G>A | p.(Arg882His) | 3 | 105 | 4064 | COSM52944 | rs147001633 |
|  | SF3B1 | NM_012433.4 | c.1997A>C | p.(Lys666Thr) | 1 | 72 | 5055 | COSM131556 | rs374250186 |
| **24767** | BCOR | NM_001123385.2 | c.573_582del | p.(Trp191Cysfs*22) | 1 | 61 | 4156 |  |  |
| **24798** | DNMT3A | NM_022552.5 | c.1628dup | p.(Arg544Profs*2) | 4 | 128 | 2888 |  |  |
| **24892** | DNMT3A | NM_022552.5 | c.2185C>T | p.(Arg729Trp) | 2 | 73 | 3047 | COSM249142 | rs200018028 |
|  | U2AF1 | NM_006758.3 | c.470A>C | p.(Gln157Pro) | 1 | 29 | 2808 | COSM211534 | rs371246226 |
|  | ASXL1 | NM_015338.6 | c.1934dup | p.(Gly646Trpfs*12) | 5 |  |  | COSM34210 |  |
| **24904** | DNMT3A | NM_022552.5 | c.2048A>G | p.(Tyr683Cys) | 6 | 118 | 1993 |  | rs780495518 |
|  | DNMT3A | NM_022552.5 | c.1586del | p.(Asp529Alafs*122) | 3 | 59 | 1872 |  |  |
|  | PPM1D | NM_003620.4 | c.1451T>G | p.(Leu484*) | 4 | 108 | 2678 | COSM7409579 |  |
|  | U2AF1 | NM_006758.3 | c.470A>G | p.(Gln157Arg) | 2 | 51 | 2297 | COSM211532 | rs371246226 |
|  | KDM6A | NM_021140.4 | c.499A>T | p.(Lys167*) | 1 | 35 | 2598 | COSM7403428 |  |
| **22344** | DNMT3A | NM_022552.5 | c.2645G>A | p.(Arg882His) | 33 | 1746 | 5225 | COSM52944 | rs147001633 |
|  | PPM1D | NM_003620.4 | c.1440del | p.(Ala481Profs*2) | 5 | 338 | 6718 | COSM6908442 |  |
| **22574** | TET2 | NM_001127208.3 | c.5374del | p.(His1792Thrfs*28) | 2 | 59 | 3883 | COSM87180 |  |
| **22622** | DNMT3A | NM_022552.5 | c.1136G>A | p.(Arg379His) | 2 | 45 | 2808 | COSM5772441 |  |
|  | TET2 | NM_001127208.3 | c.2079dup | p.(Leu694Thrfs*18) | 6 | 425 | 7155 |  |  |
| **22736** | DNMT3A | NM_022552.5 | c.994G>A | p.(Gly332Arg) | 1 | 22 | 1509 | COSM2270955 | rs760854242 |
|  | CALR | NM_004343.4 | c.1194G>T | p.(Glu398Asp) | 2 | 15 | 769 | COSM1738023 |  |
| **22786** | TET2 | NM_001127208.3 | c.4393C>T | p.(Arg1465*) | 1 | 71 | 5030 | COSM42016 |  |
|  | TET2 | NM_001127208.3 | c.4546C>T | p.(Arg1516*) | 2 | 56 | 2582 | COSM43420 | rs370735654 |
|  | TET2 | NM_001127208.3 | c.4894C>T | p.(Gln1632*) | 3 | 135 | 4124 | COSM3719314 |  |
|  | SMC1A | NM_006306.4 | c.34T>G | p.(Phe12Val) | 3 | 65 | 2187 |  |  |
| **23927** | DNMT3A | NM_022552.5 | c.2249C>G | p.(Pro750Arg) | 1 | 93 | 6664 |  |  |
|  | TET2 | NM_001127208.3 | c.2375C>G | p.(Ser792*) | 3 | 289 | 8528 |  |  |
|  | CBL | NM_005188.4 | c.1211G>A | p.(Cys404Tyr) | 1 | 48 | 4475 | COSM34068 | rs192712314 |
| **24149** | DNMT3A | NM_022552.5 | c.2194T>A | p.(Phe732Ile) | 1 | 74 | 5220 |  | rs149043640 |
| **24277** | TET2 | NM_001127208.3 | c.5565_5593del | p.(Asp1856Serfs*9) | 1 | 24 | 2111 |  |  |
| **24530** | TET2 | NM_001127208.3 | c.2884C>T | p.(Gln962*) | 5 | 313 | 6338 |  | rs745887194 |
| **24537** | DNMT3A | NM_022552.5 | c.2732G>A | p.(Cys911Tyr) | 1 | 17 | 1277 | COSM9277296 | rs906113912 |
| **24704** | DNMT3A | NM_022552.5 | c.2339T>C | p.(Ile780Thr) | 2 | 65 | 3576 | COSM1583121 | rs370751539 |
| **22850** | STAG2 | NM_001042749.2 | c.3306G>C | p.(Met1102Ile) | 2 | 12 | 484 |  |  |
| **23128** | DNMT3A | NM_022552.5 | c.901C>T | p.(Arg301Trp) | 1 | 77 | 6962 | COSM2911819 |  |
| **23167** | CBL | NM_005188.4 | c.1259G>T | p.(Arg420Leu) | 1 | 62 | 4990 | COSM34080 |  |
| **23178** | GATA1 | NM_002049.4 | c.77C>G | p.(Ser26Cys) | 2 | 77 | 4938 |  |  |
| **24028** | TP53 | NM_001126112.3 | c.818G>A | p.(Arg273His) | 1 | 100 | 6868 | COSM10660 | rs28934576 |
|  | PPM1D | NM_003620.4 | c.1354del | p.(Val452Phefs*5) | 1 | 42 | 3516 |  |  |
| **24044** | TET2 | NM_001127208.3 | c.4138C>T | p.(His1380Tyr) | 1 | 10 | 735 | COSM87161 | rs577810844 |
|  | KRAS | NM_033360.4 | c.35G>T | p.(Gly12Val) | 1 | 47 | 3654 | COSM520 | rs121913529 |
|  | KDM6A | NM_021140.4 | c.2968G>T | p.(Val990Leu) | 4 | 19 | 470 |  |  |

**Supplementary Table S5: comparison of characteristics of MI(+) subjects depending on their CHIP and mLOY status**

|  | CHIP(-) subjects  n=70 (47%) | CHIP(+) subjects  n=79 (53%) | p-value | mLOY(-) subjects  n=53 (55%) | mLOY(+) subjects  n=44 (45%) | p-value |
| --- | --- | --- | --- | --- | --- | --- |
| Male, n (%) | 49 (70%) | 49 (62%) | 0.387 | - | - | - |
| Age (years), median (Q1;Q3) | 81 (77;84) | 83 (80;87) | **0.011** | 80 (77;84) | 82 (79;85) | 0.209 |
| *Risk factors* | | | | | | |
| BMI (kg/m^2^), median (Q1;Q3) | 25.7 (23.8;29.1) | 25.1 (22.5;28.3) | 0.180 | 25.3 (23.1;28.4) | 25.3 (23.9;28.5) | 0.437 |
| Diabetes, n (%) | 29 (41%) | 18 (23%) | **0.023** | 19 (36%) | 14 (33%) | 0.807 |
| Hypertension, n (%) | 50 (75%) | 57 (78%) | 0.985 | 36 (69%) | 32 (76%) | 0.615 |
| Cholesterolemia (g/L), median (Q1;Q3) | 1.4 (1.3;1.7) | 1.5 (1.2;1.7) | 0.695 | 1.4 (1.3;1.7) | 1.4 (1.1;1.6) | 0.148 |
| LDL-c (g/L), median (Q1;Q3) | 0.76 (0.61;0.87) | 0.80 (0.63;1.05) | 0.409 | 0.77 (0.66:1.03) | 0.71 (0.53;0.86) | 0.085 |
| Smoking, n (%) | 4 (8.3%) | 2 (3.2%) | 0.399 | 3 (6.8%) | 1 (3.1%) | 0.634 |
| *Blood test* | | | | | | |
| Hemoglobin (g/dL), median (Q1;Q3) | 13.4 (12.6;14.4) | 12.9 (11.8;14.0) | 0.409 | 13.6 (12.6;14.5) | 13.1 (12.2;14.4) | 0.572 |
| White blood cells (G/L), median (Q1;Q3) | 6.1 (5.1;7.5) | 6.6 (5.7;7.4) | 0.125 | 6.6 (5.4;7.7) | 6.6 (5.7;7.4) | 0.793 |
| Neutrophils (G/L), median (Q1;Q3) | 3.6 (2.7;4.5) | 4.0 (3.3;4.6) | 0.085 | 4.0 (3.1;5.1) | 4.1 (3.4;4.7) | 0.828 |
| Platelets (G/L), median (Q1;Q3) | 185 (155;222) | 215 (184:253) | **0.007** | 195 (172;227) | 200 (167;231) | 0.642 |

For statistical analysis, logistical regression (adjusted on age and sex) and/or Fisher’s test were used to compare qualitative variables and linear regression (adjusted on age and sex) and/or ANOVA were used to compare quantitative variables.

**Supplementary Table S6: comparison of characteristics of MI(-) subjects depending on their CHIP and mLOY status**

|  | CHIP(-) subjects  n=175 (59%) | CHIP(+)subjects  n=122 (41%) | p-value | mLOY(-) subjects  n=84 (68%) | mLOY(+) subjects  n=39 (32%) | p-value |
| --- | --- | --- | --- | --- | --- | --- |
| Male, n (%) | 99 (57%) | 60 (49%) | 0.237 | - | - | - |
| Age (years), median (Q1;Q3) | 73 (70;77) | 74 (71;79) | **0.030** | 72 (70;76) | 75 (71;78) | **0.004** |
| *Risk factors* | | | | | | |
| BMI (kg/m^2^), median (Q1;Q3) | 25.8 (23.8;28.2) | 25.1 (22.9;27.8) | 0.294 | 26.0 (24.3;28.3) | 25.9 (24.1;28.1) | 0.445 |
| Diabetes, n (%) | 34 (20%) | 13 (11%) | 0.055 | 13 (16%) | 2 (5%) | 0.099 |
| Hypertension, n (%) | 152 (87%) | 103 (84%) | 0.397 | 73 (87%) | 32 (82%) | 0.483 |
| Cholesterolemia (g/L), median (Q1;Q3) | 2.2 (2.0;2.4) | 2.2 (2.0;2.5) | 0.915 | 2.2 (2.0;2.3) | 2.2 (2.1;2.4) | 0.263 |
| LDL-c (g/L), median (Q1;Q3) | 1.4 (1.2;1.6) | 1.4 (1.2;1.6) | 0.719 | 1.4 (1.2;1.5) | 1.5 (1.3;1.6) | 0.120 |
| Smoking, n (%) | 17 (9.7%) | 9 (7.4%) | 0.537 | 8 (9.5%) | 8 (20.5%) | 0.149 |

For statistical analysis, logistical regression (adjusted on age and sex) and/or Fisher’s test were used to compare qualitative variables and linear regression (adjusted on age and sex) and/or ANOVA were used to compare quantitative variables.

Blood counts were not available for MI(-) subjects

**Supplementary Table S7: CHIP associated with *DNMT3A* or *TET2* mutations do not present differential effect on inflammation or atherosclerotic burden**

|  | CHIP (-) | *DNMT3A* | *TET2* | Other CHIP |
| --- | --- | --- | --- | --- |
| Inflammation (all subjects, n = 446) | | | | |
| Subjects, n | 245 | 97 | 61 | 61 |
| hsCRP (mg/L), median (Q1;Q3) | 1.64 (1.00;3.69) | 2.00 (1.16;3.01) | 2.38 (1.00:4.00) | 1.42 (1.00;4.00) |
| hsCRP > 2 mg/L, n (%) | 61 (44.2) | 27 (45.8) | 17 (51.5) | 16 (43.2) |
| Atherosclerosis burden evaluation in MI(+) subjects | | | | |
| Multitroncular lesions, n (%) | 29 (42.6) | 19 (47.5) | 12 (48.0) | 11 (55.0) |
| Carotid stenosis ≥ 50%, n (%) | 2 (3.1) | 0 | 3 (12.0) | 2 (10.0) |
| Global atheroma volume (mm^3^), median (Q1;Q3) | 455.0 (374.0;555.0) | 530.0 (402.0;603.0) | 522.5 (398.8:645.5) | 515.0 (492.0;520.0) |
| Atherosclerosis burden evaluation in MI(-) subjects | | | | |
| Patients with atherosclerotic plaks, n (%) | 81 (59.6) | 23 (54.8) | 15 (60.0) | 20 (66.7) |
| Number of plaque, median (Q1;Q3) | 2 (1;2) | 2 (1;2) | 2 (1;2) | 1 (1;2) |
| Intima-Media Thickness, median (Q1;Q3) | 0.672 (0.605;0.760) | 0.677 (0.586;0.732) | 0.700 (0.625;0.820) | 0.677 (0.610;0.801) |
| Cardiovascular events in MI(-) subjects | | | | |
| MI incidence, n (%) | 45 (25.7) | 19 (33.9) | 9 (25.7) | 9 (21.9) |

Data are expressed as numbers and frequency or median, first and third quartiles. For quantitative values, comparisons were made by linear regression of log values adjusted for age and sex. For qualitative parameters, comparisons were made by the fisher test.

**Supplementary Table S8: Important clones of CHIP and mLOY are not associated with increased inflammation, atherosclerosis or CVE**

|  | CHIP | | | mLOY | | | |
| --- | --- | --- | --- | --- | --- | --- | --- |
|  | CHIP (-)  n = 245 | VAF 2-5%  n = 113 | VAF≥5%  n = 88 | | mLOY (-)  n=129 | mLOY <50%  n=81 | mLOY ≥50%  n=10 |
| Inflammation | | | | | | | |
| hsCRP (mg/L), median (Q1;Q3) | 1.64 (1.00;3.69) | 1.85 (1.00;3.00) | 2.00 (1.13;4.43) | | 1.4 (0.97;2.54) | 1.85 (1.00;4.00) | 5.59 (1.37;7.92) |
| hsCRP > 2 mg/L, n (%) | 61 (44.2) | 27 (42.9) | 26 (47.3) | | 28 (33.7) | 25 (44.6) | 4 (57.1) |
| Atherosclerosis burden evaluation in MI(+) patients | | | | | | | |
| Subjects (n) | 70 | 49 | 30 | | 46 | 46 | 5 |
| Multitroncular lesions, n (%) | 29 (42.6) | 21 (42.8) | 18 (62.1) | | 21 (46.7) | 23 (51.1) | 1 (20.0) |
| Carotid stenosis ≥ 50%, n (%) | 2 (3.1) | 3 (6.4) | 2 (6.7) | | 2 (4.4) | 3 (6.7) | 0 |
| Global atheroma volume (mm^3^) , median (Q1;Q3) | 455.0 (374.0;555.0) | 506.0 (398.8;608.2) | 530.0 (414.0;661.0) | | 618.0 (463.0;723.0) | 473.5 (394.5;599.2) | 347.0  (NA) |
| Atherosclerosis burden evaluation in MI(-) subjects | | | | | | | |
| Subjects (n) | 175 | 64 | 58 | | 83 | 35 | 5 |
| Patients with atherosclerotic plaque, n (%) | 81 (59.6) | 31 (64.6) | 23 (57.5) | | 34 (55.7) | 15 (60) | 4 (100) |
| Number of plaque, median (Q1;Q3) | 2 (1;2) | 1 (1;2) | 1 (1;2) | | 2 (1;2) | 2 (1;2) | 2 (1.75;2) |
| Intima-Media Thickness (mm), median (Q1;Q3) | 0.672 (0.605;0.760) | 0.675 (0.595;0.760) | 0.697 (0.602;0.740) | | 0.677 (0.619;0.764) | 0.720 (0.600;0.830) | 0.560 (0.541;0.626) |
| Cardiovascular events in MI(+) subjects | | | | | | | |
| Subjects (n) | 70 | 49 | 30 | | 46 | 46 | 5 |
| MI recurrence at 1 year, n (%) | 2 (2.85) | 0 (0) | 1 (3.3) | | 2 (4.3) | 1 (2.2) | 0 (0.0) |

Data are expressed as numbers and frequency or median, first and third quartiles. For quantitative values, comparisons were made by linear regression of log values adjusted for age and sex. For qualitative parameters, comparisons were made by the fisher test and logistic regression. For each variable, results are expressed among patients with available value.

**Supplementary Table 9: mLOY do not impact the atherosclerotic burden in the presence of CHIP**

|  | CHIP (-)  mLOY (-) | CHIP (+)  mLOY (-) | CHIP (-)  mLOY (+) | CHIP (+)  mLOY (+) | p-value |
| --- | --- | --- | --- | --- | --- |
| Atherosclerosis burden evaluation in MI(+) subjects | | | | | |
| Males, n | 27 | 26 | 22 | 22 | - |
| Multitroncular lesions, n (%) | 13 (48.1) | 12 (48.0) | 8 (38.1) | 12 (54.5) | 0.63 |
| Carotid stenosis ≥ 50%, n (%) | 1 (3.8) | 1 (3.8) | 1 (4.5) | 2 (9.5) | 0.8 |
| Global atheroma volume, median (Q1;Q3) | 412.0 (411.0;705.0) | 609.5 (491.5;720.5) | 455.0 (389.0;529.0) | 511.0 (345.8;599.8) | 0.81 |
| Atherosclerosis burden evaluation in MI(-) subjects | | | | | |
| Males, n | 51 | 33 | 22 | 17 | - |
| Patients with atherosclerotic plaque, n (%) | 19 (37.2) | 15 (45.4) | 9 (40.9) | 10 (58.8) | 0.19 |
| Number of plaque, median (Q1;Q3) | 2 (1;2) | 1 (1;2) | 1 (1;2) | 2 (1.25;2) | 0.50 |
| Intima-Media Thickness, median (Q1;Q3) | 0.745 (0.676;0.850) | 0.675 (0.597;0.745) | 0.720 (0.580;0.795) | 0.682 (0.570;0.856) | 0.56 |

Data are expressed as numbers and frequency or median, first and third quartiles. For quantitative values, comparisons were made by linear regression of log values adjusted for age and sex. For qualitative parameters, comparisons were made by the fisher test. For each variable, results are expressed among patients with available value.

**Supplementary Table S10: Characteristics of MI(-) patients at the time of inclusion depending on whether they presented a MI during follow up or not**

|  | MI(-) subjects  with MI during FU  n = 79 | MI(-) subjects  without MI during FU  n = 218 | p-value |
| --- | --- | --- | --- |
| Male, n (%) | 41 (51.9) | 118 (54.1) | 0.793 |
| Median age, years (Q1;Q3) | 74.4 (71.2;78.3) | 73.3 (70.5;77.4) | 0.069 |
| Cardiovascular risk factors | | | |
| BMI (kg/m^2^), median (Q1;Q3) | 25.4 (23.3;27.1) | 25.6 (23.7;28.4) | 0.196 |
| Diabetes, n (%) | 7 (8.9) | 40 (18.5) | 0.065 |
| Hypertension, n (%) | 66 (83.5) | 189 (86.7) | 0.367 |
| Total cholesterol (g/L), median (Q1;Q3) | 2.30 (2.07;2.52) | 2.21 (1.98;2.44) | 0.197 |
| LDL-c (g/L), median (Q1;Q3) | 1.39 (1.24;1.64) | 1.39 (1.17;1.58) | 0.373 |
| HDL-c (g/L), median (Q1;Q3) | 0.60 (0.50;0.74) | 0.60 (0.49;0.68) | 0.127 |
| Smoking, n (%) | 10 (12.7) | 16 (7.3) | 0.142 |
| Prevalence of CHIP and mLOY | | | |
| CHIP prevalence, n (%) | 34 (43.0) | 88 (40.4) | 0.860 |
| CHIP prevalence with VAF ≥5%, n (%) | 17 (21.5) | 41 (18.8) | 0.842 |
| Males tested for mLOY, n | 31 | 92 | - |
| mLOY prevalence, n (%) | 12 (38.7) | 27 (29.3) | 0.478 |
| Prevalence of mLOY involving ≥50% cells, n (%) | 0 (0) | 5 (5.4%) | 0.210 |
| CHIP(+)/mLOY(+), n (%) | 6 (19.3) | 11 (11.9) | 0.47 |

Data are expressed as numbers and frequency or median, first and third quartiles. For quantitative values, comparisons were made by linear regression of log values adjusted for age and sex. For qualitative parameters, comparisons were made by the fisher test and logistic regression. For each variable, results are expressed among patients with available value.

**Supplementary Table S11: CHIP may accelerate the incidence MI in the absence of mLOY in male subjects**

|  | CHIP (-) | CHIP (+) | p-value | mLOY (-) | mLOY (+) | p-value | CHIP (-)  mLOY (-) | CHIP (+)  mLOY (-) | CHIP (-)  mLOY (+) | CHIP (+)  mLOY (+) | p-value |
| --- | --- | --- | --- | --- | --- | --- | --- | --- | --- | --- | --- |
| MI(+) subjects | | | | | | | | | | | |
| Subjects, n | 70 | 79 | - | 53 | 44 | - | 27 | 26 | 22 | 22 | - |
| MI recurrence at 1 year, n (%) | 2 (2.9%) | 1 (1.3%) | 0.60 | 1 (1.9%) | 1 (2.3%) | 0.99 | 1 (3.7%) | 0 | 0 | 1 (4.5%) | 0.564 |
| MI(-) subjects | | | | | | | | | | | |
| Subjects, n | 175 | 122 | - | 84 | 39 | - | 51 | 33 | 22 | 17 | - |
| MI incidence, n (%) | 45 (25.7) | 34 (27.9) | 0.957 | 19 (22.6) | 12 (30.8) | 0.464 | 9 (17.6) | 10 (30.3) | 6 (27.3) | 6 (35.3) | 0.4 |
| Time to MI, median (Q1;Q3) | 5.299 (2.787;  7.843) | 4.404 (2.550;  6.294) | 0.2256 | 3.47 (1.879;  6.313) | 6.00 (3.655;  8.045) | 0.1031 | 6.313 (4.626;  8.246) | 2.511 (1.429;  3.709) | 7.360 (3.874;  9.845) | 5.288 (3.019;  6.593) | **0.0104** |

Data are expressed as numbers and frequency or median, first and third quartiles. For the “time to MI” in MI(-) subjects, comparisons were performed by Mann&Whitney test for CHIP and mLOY separately and Kruskall-Wallis test for combinations of CHIP+/-mLOY. For qualitative parameters, comparisons were made by the fisher test. For each variable, results are expressed among patients with available value.
